# Transmission of SARS-CoV-2 Lineage B.1.1.7 in England: Insights from linking epidemiological and genetic data

**DOI:** 10.1101/2020.12.30.20249034

**Authors:** Erik Volz, Swapnil Mishra, Meera Chand, Jeffrey C. Barrett, Robert Johnson, Lily Geidelberg, Wes R Hinsley, Daniel J Laydon, Gavin Dabrera, Áine O’Toole, Roberto Amato, Manon Ragonnet-Cronin, Ian Harrison, Ben Jackson, Cristina V. Ariani, Olivia Boyd, Nicholas J Loman, John T McCrone, Sónia Gonçalves, David Jorgensen, Richard Myers, Verity Hill, David K. Jackson, Katy Gaythorpe, Natalie Groves, John Sillitoe, Dominic P. Kwiatkowski, The COVID-19 Genomics UK (COG-UK) consortium, Seth Flaxman, Oliver Ratmann, Samir Bhatt, Susan Hopkins, Axel Gandy, Andrew Rambaut, Neil M Ferguson

## Abstract

The SARS-CoV-2 lineage B.1.1.7, now designated Variant of Concern 202012/01 (VOC) by Public Health England, originated in the UK in late Summer to early Autumn 2020. We examine epidemiological evidence for this VOC having a transmission advantage from several perspectives. First, whole genome sequence data collected from community-based diagnostic testing provides an indication of changing prevalence of different genetic variants through time. Phylodynamic modelling additionally indicates that genetic diversity of this lineage has changed in a manner consistent with exponential growth. Second, we find that changes in VOC frequency inferred from genetic data correspond closely to changes inferred by S-gene target failures (SGTF) in community-based diagnostic PCR testing. Third, we examine growth trends in SGTF and non-SGTF case numbers at local area level across England, and show that the VOC has higher transmissibility than non-VOC lineages, even if the VOC has a different latent period or generation time. Available SGTF data indicate a shift in the age composition of reported cases, with a larger share of under 20 year olds among reported VOC than non-VOC cases. Fourth, we assess the association of VOC frequency with independent estimates of the overall SARS-CoV-2 reproduction number through time. Finally, we fit a semi-mechanistic model directly to local VOC and non-VOC case incidence to estimate the reproduction numbers over time for each. There is a consensus among all analyses that the VOC has a substantial transmission advantage, with the estimated difference in reproduction numbers between VOC and non-VOC ranging between 0.4 and 0.7, and the ratio of reproduction numbers varying between 1.4 and 1.8. We note that these estimates of transmission advantage apply to a period where high levels of social distancing were in place in England; extrapolation to other transmission contexts therefore requires caution.

## Introduction

A novel SARS-CoV-2 lineage, originally termed variant B.1.1.7, is rapidly expanding its geographic range and frequency in England. The lineage was detected in November 2020, and likely originated in September 2020 in the South East region of England. As of 20 December 2020, the regions in England with the largest numbers of confirmed cases of the variant are London, the South East, and the East of England. The variant possesses a large number of non-synonymous substitutions of immunologic significance^1^. The N501Y replacement on the spike protein has been shown to increase ACE2 binding ^2,3^ and cell infectivity in animal models^4^, while the P618H replacement on the spike proteins adjoins the furin-cleavage site^5^. The variant also possesses a deletion at positions 69 and 70 of the spike protein (Δ69-70) which has been associated with diagnostic test failure for the ThermoFisher TaqPath probe targeting the spike protein^6^. Whilst other variants with Δ69-70 are also circulating in the UK, the absence of detection of the S gene target in an otherwise positive PCR test increasingly appears to be a highly specific marker for the B.1.1.7 lineage. Surveillance data from national community testing (“Pillar 2”) showed a rapid increase in S-gene target failures (SGTF) in PCR testing for SARS-CoV-2 in November and December 2020, and the B.1.1.7 lineage has now been designated Variant of Concern (VOC) 202012/01 by Public Health England (PHE).

Phylogenetic studies carried out by the UK COVID-19 Genomics Consortium (COG-UK)^7^ provided the first indication that the VOC has an unusual accumulation of substitutions and was growing at a large rate relative to other circulating lineages. Here we analyse VOC whole genomes collected between October and 5 December 2020 and find that the rate of increase in the frequency of VOC is consistent with a transmission advantage over other circulating lineages in the UK. To substantiate these findings, we investigate time trends in the proportion of PCR tests exhibiting SGTF across the UK on ∼275,000 test results as a biomarker of VOC infection, and examine the relationship between local epidemic growth and the frequency of the VOC. We demonstrate that increasing reproduction numbers (‘R’ values) are associated with increased SGTF frequency among reported cases, our biomarker of VOC infection, and confirm this association through a variety of analytical approaches. Critically, we find evidence that non-pharmaceutical interventions (NPIs) were sufficient to control non-VOC lineages to reproduction numbers below 1 during the November 2020 lockdown in England, but that at the same time the NPIs were insufficient to control the VOC.

### Origins and expansion of VOC 202012/01

We examined the time and location of sampling of 1,904 VOC whole genomes collected between October and 5 December 2020, combined with a genetic background of 48,128 genomes collected over the same period. Sequences of the VOC were widely distributed across 199 lower tier local authorities (LTLAs) in England, but highly concentrated in the South East (n=875), London (n=636) and East of England (n=293). Relative to this genetic background, the growth of the VOC lineage is consistent with it having a selective advantage over circulating SARS-CoV-2 variants in England (Figure 1A). While rapid growth of the variant was first observed in the South East, similar growth patterns are observed later in London, East of England, and now more generally across England. Across these regions, we estimate similar growth differences between the VOC and non-VOC lineages of +49% to 53% per generation (Supporting Table S1) by fitting a logistic growth model to the frequency of VOC sequence samples through time and adjusting for an approximate mean generation time of SARS-CoV-2 of 6.5 days (see Supporting Methods) ^8,9^.

**Figure 1.**
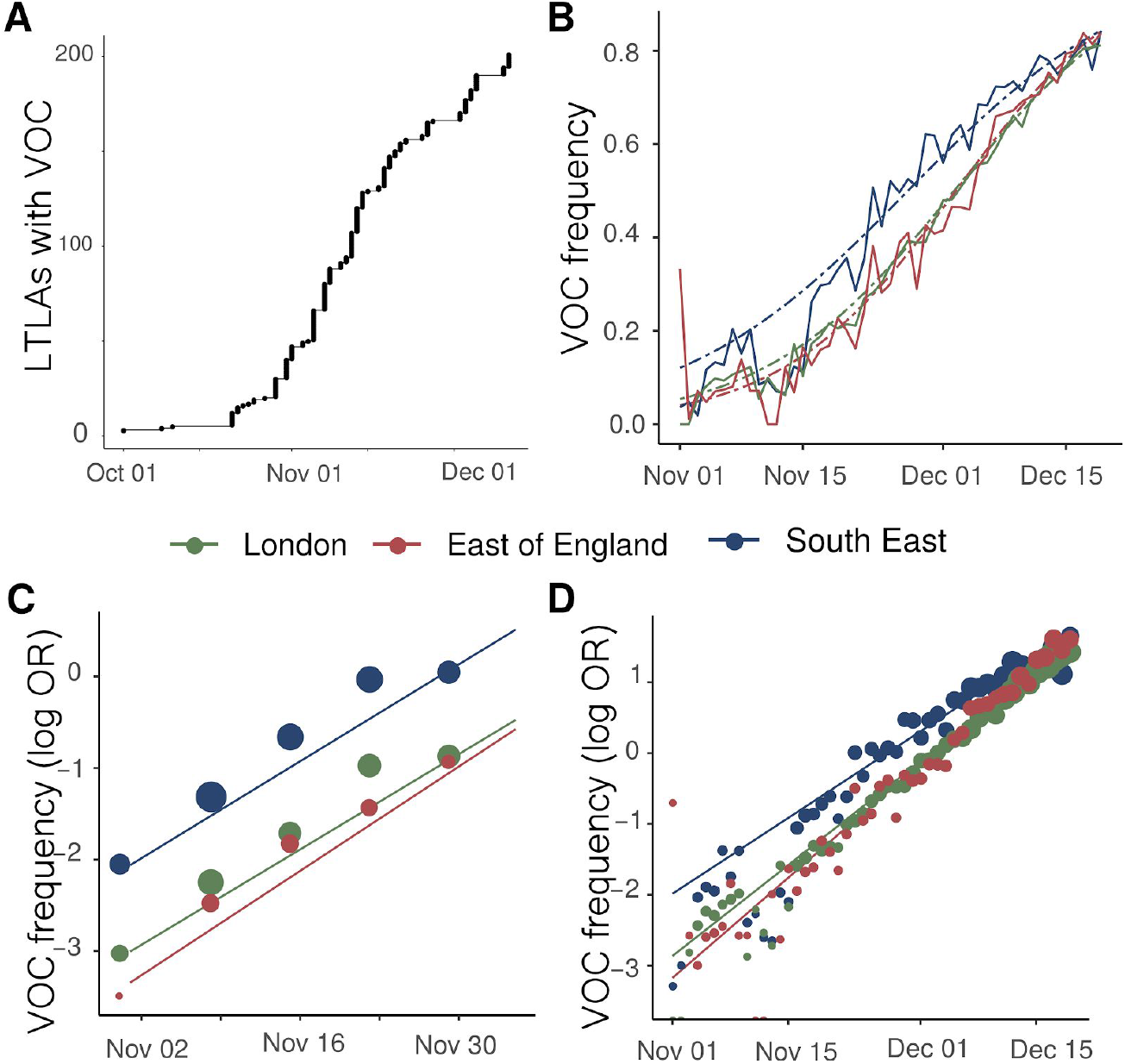
Expansion and growth of the VOC *202012/01* lineage. A) The number of UK LTLAs reporting at least one sampled VOC genome. B) Empirical (solid) and estimated (dash) frequency of TPR-adjusted SGTF in three regions of England. C) Empirical (points) and estimated (line) frequency (log odds) of VOC inferred from genomic data by epidemiological week. D) Empirical (points) and estimated (line) frequency (log odds) of SGTF based on the same data as B.

### S gene target failure in SARS-CoV-2 testing as a biomarker for the VOC

The UK has a high throughput national testing system for community cases, based in a small number of large laboratories. We were able to extend our genomic analyses to epidemiologic case data, because the VOC lineage is not detected in the S-gene target in an otherwise positive PCR test (ThermoFisher TaqPath as performed in the UK national testing system). Several SARS-CoV-2 variants can result in SGTF, but since mid-November, more than 97% of Pillar 2 PCR tests showing SGTF are due to the VOC lineage^10^. Before mid-November 2020, the frequency of SGTF among PCR positives was a poorer proxy for frequency of the VOC. We therefore developed a Gaussian Markov Random Field model (see Supplementary Information, Figure S1) to predict the proportion of SGTF cases attributable to the VOC lineage by area and week, here termed the true positive rate (TPR), and the number of SGTF cases attributable to the VOC. In turn, the corresponding false-positives were attributed to the S-gene positive case (S+) category.

### Trends in SARS-CoV-2 cases with S gene target failure that are attributed to the VOC

SGTF data were available for 35% of Pillar 2 positive test results between November 26 to December 13, 2020. Given the greater abundance of SGTF data, a more detailed picture of the VOC frequency over time can be discerned after our TPR adjustments. Overall, empirical and estimated frequencies of TPR-adjusted SGTF cases show a similar pattern of expansion as frequencies estimated from genetic data in terms of time, region, and rate of growth (Figure 1D). As of December 13, SGTF is detected in all regions of England (Figure S2), and the estimated frequency of TPR-adjusted SGTF ranges from 15% in Yorkshire and the Humber to 85% in the South East, where the VOC was first detected. Changes in COVID-19 infections correlate with raw (not adjusted for TPR) SGTF cases on a regional basis. Figures 2 and S3 shows the time trends of SGTF (S-) cases, S-gene positive cases (S+) and total PCR positive cases by NHS England Sustainability and Transformation Plan (STP) areas (a geographic subdivision of NHS Regions). Visually, it is clear that while lockdown successfully controlled S+ cases in virtually every STP, S-case numbers increased during lockdown.

**Figure 2.**
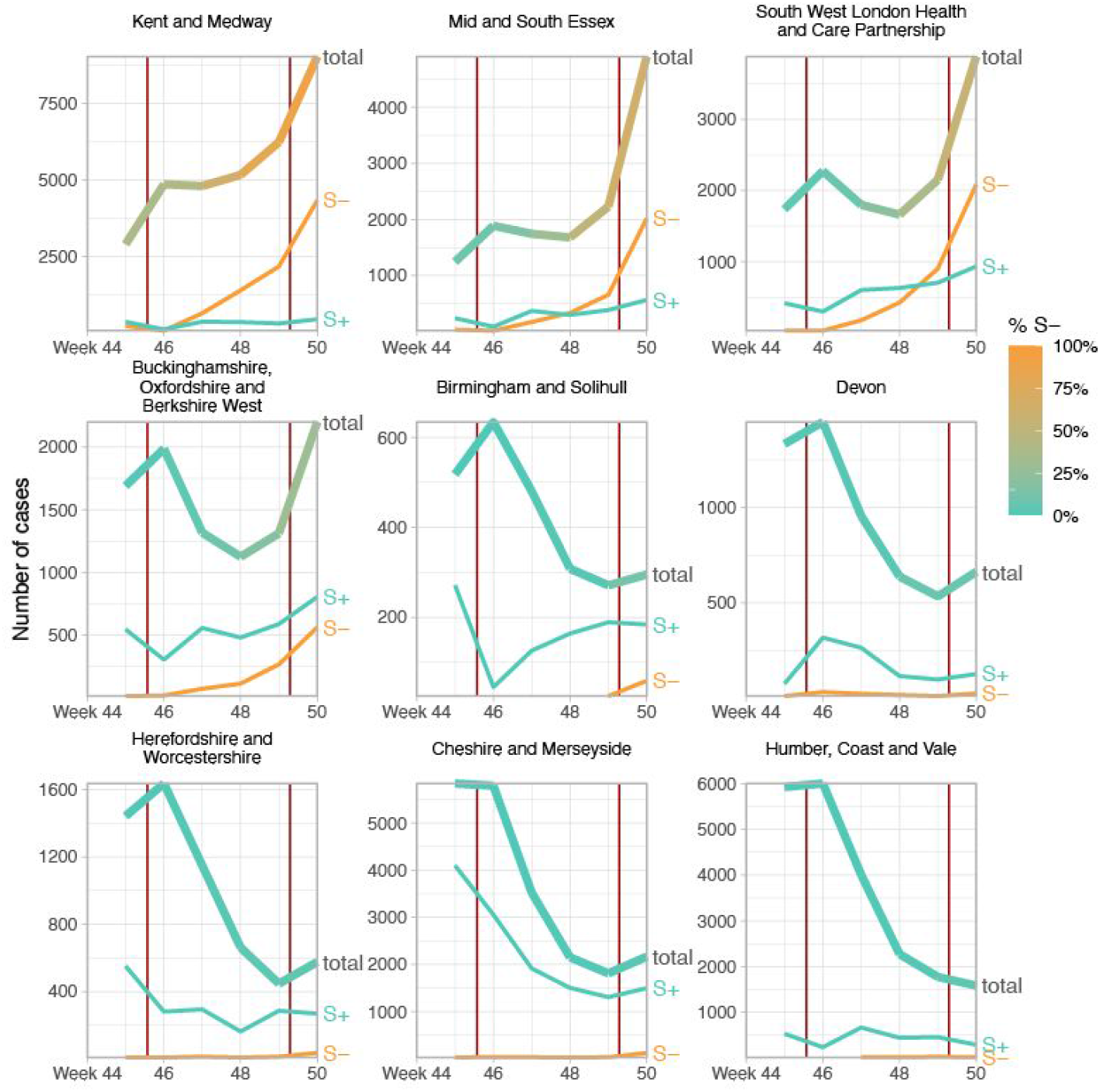
Case trends in a subset of NHS STP areas. Total cases reported are shown as a thick line. A subset of these - those tested in the 3 largest “Lighthouse” laboratories - were tested for SGTF. The total cases line is coloured according to percentage S-among those tested. Counts of S+ and S-reported via the PHE SGSS system are shown by the thin lines. The dates of the second lockdown are indicated by the vertical red lines. Nine representative NHS STP areas from all regions of England are ordered by decreasing percentage S-in the most recent week of data. Raw SGTF data are shown here (not adjusted for TPR), so S-cases in earlier weeks include other non-VOC lineages, especially outside the East and South East of England. Plots for all STP areas are shown in Figure S3.

### Transmission advantage of the VOC

To examine the differences between S- and S+ growth rates, we focus on epidemiological weeks 46-50 (8th November-12th December). We estimate the total S- and S+ in each STP and week by adjusting counts upwards in proportion to total cases reported in each STP and week. We then calculate the week on week growth factor in both S- and S+ cases by dividing the case numbers in week *t+1* by the case numbers in week *t*. Given an assumed mean generation time of SARS-CoV-2 of 6.5 days^9^, we correct these weekly growth factors by raising them to the power of 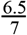 to ensure they can be interpreted as approximate reproduction numbers. For each STP and week, we compute both the ratio and difference of the resulting empirical reproduction number of the S-negative cases to that of the S-positive cases (Figure 3). Overall, the median multiplicative advantage is 1.74 for the VOC, and the median additive advantage is 0.63, showing a clear advantage of the VOC for both metrics.

**Figure 3.**
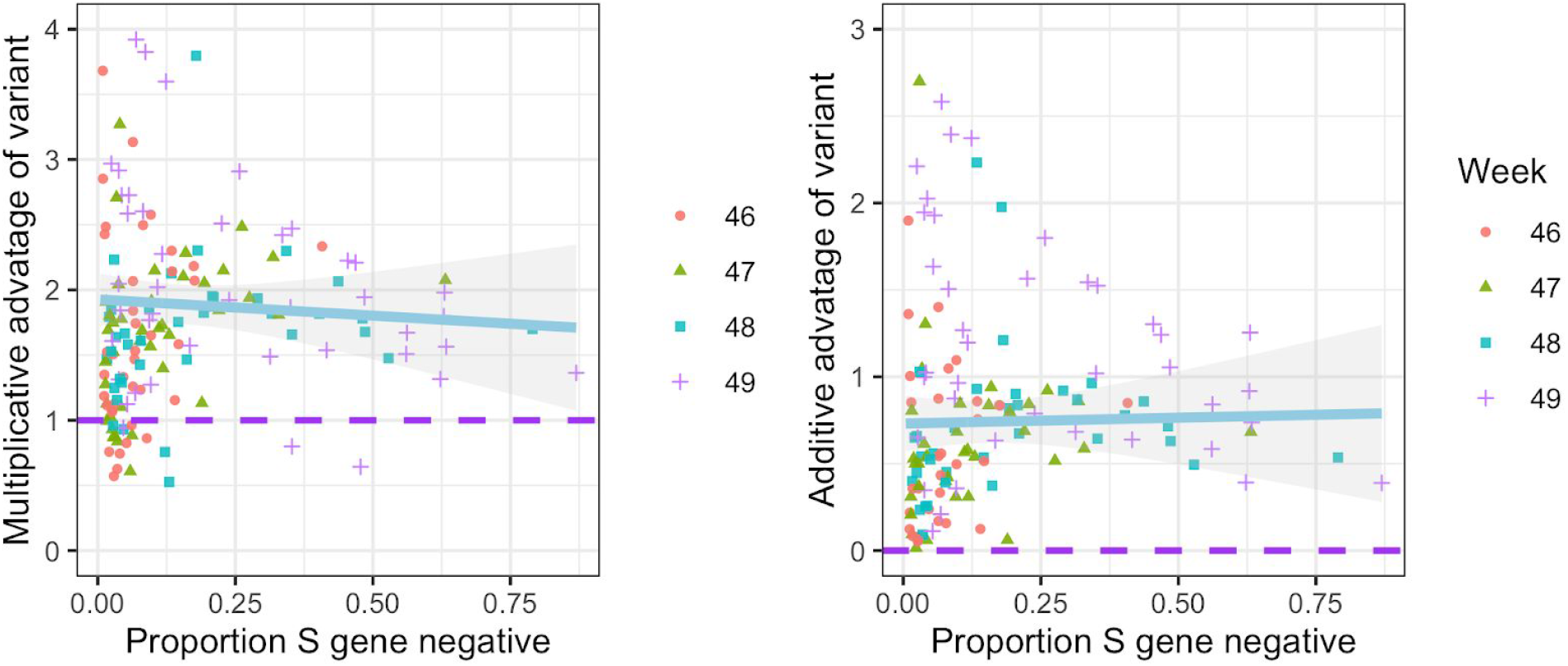
Empirical data analysis of the advantage in weekly growth factors (cases in week t+1 divided by cases in week t) for the VOC versus non-VOC lineages. Each point represents either the ratio (left) or difference (right) of weekly growth factors for the VOC versus non-variant for an NHS England STP area and week, using the raw SGTF data shown in Figure S1 (not correcting for TPR). Colours and shapes differentiate epi weeks. Numbers above 1 on the top plot and above 0 on the bottom plot show a transmission advantage. The blue line represents the mean advantage for a particular proportion of VOC among all cases, and the grey lines the 95% envelope. Scatter at low frequencies largely reflects statistical noise due to low counts.

### Paired growth rate trends of the VOC and non-VOC lineages demonstrate an increase in the reproduction number

We next tested the hypothesis that the higher growth rates of the VOC compared to other circulating lineages might be due solely to shorter generation times (e.g. a shorter incubation period), rather than increased transmissibility (R). To this end, we compared the number of NHS STP areas in which both VOC and non-VOC cases increased or decreased (Table 1). If the VOC had the same reproduction number as non-VOC but a shorter generation time, VOC cases are expected to grow faster than non-VOC cases in areas where non-VOC grew. However VOC cases are expected to *decline* faster than non-VOC cases where non-VOC declined. Furthermore, areas where VOC grew but non-VOC declined would, on average, be equally balanced by areas where the opposite was true. That is, if only the generation interval of the VOC had shortened, the proportion of areas with positive growth of the VOC and negative growth of the non-VOC would be highly correlated with the proportion of areas with negative growth of the VOC and positive growth of the non-VOC. However, of 168 STP-weeks (42 STP areas, weekly growth factors for weeks 46-49) there were 97 STP-weeks where growth was observed in S- and decline was observed in S+, but only 1 STP-week where the opposite was true (Table 1), indicating strong evidence against S+ and S-reproduction numbers being equal (McNemar’s Chi-square test with continuity correction test statistic 92.02, *p* < 1e-15). Comparing the empirical distribution of growth factors from S+ and S-with the nonparametric Kolmogorov–Smirnov test results in rejecting the null hypothesis (*p* < 1e-15) that the two arise from the same probability distribution.

**Table 1.**
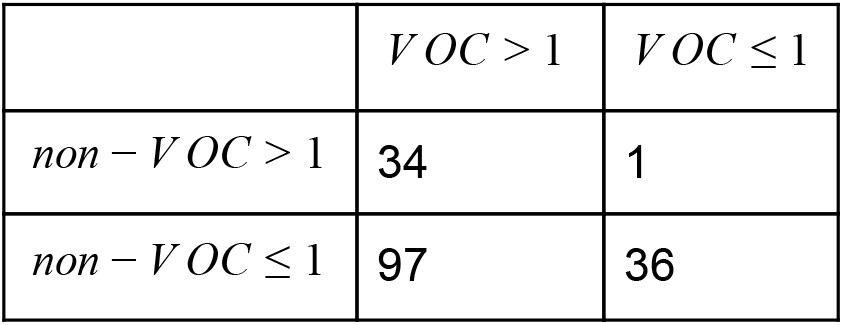
Contingency table of VOC and non-VOC weekly growth factors derived from raw SGTF data within 42 NHS STP areas for weeks 46-49, stratified by increasing (>1) and declining incidence(≤ 1). The imbalance in off-diagonal elements gives strong evidence of increased transmissibility, even if the VOC had an altered generation time distribution.

### Share of age groups among VOC and non-VOC cases

To assess differences in the age distribution of VOC versus non-VOC cases, we considered S- and S+ case numbers in weeks 46-51 across NHS STP regions. Case numbers were standardised for differences in the population age composition in each area, weighted to compare S-cases from each NHS STP region and each epidemiological week with an equal number of S+ cases from that same STP and week (a case-control design), and aggregated over STP weeks. Accounting for binomial sampling variation and variation by area and week, we observe significantly more S-cases, our biomarker of VOC cases, among individuals aged 0-19 as compared to S+ cases, and significantly fewer S-cases among individuals aged 60-79 (Figure 4). This trend is seen in each of the regions of England most affected by the VOC thus far (East of England, London, South East and Midlands), and similar differences are seen between the raw (non-case control weighted, and non-age-standardised) age distributions of S+ and S-cases.

**Figure 4.**
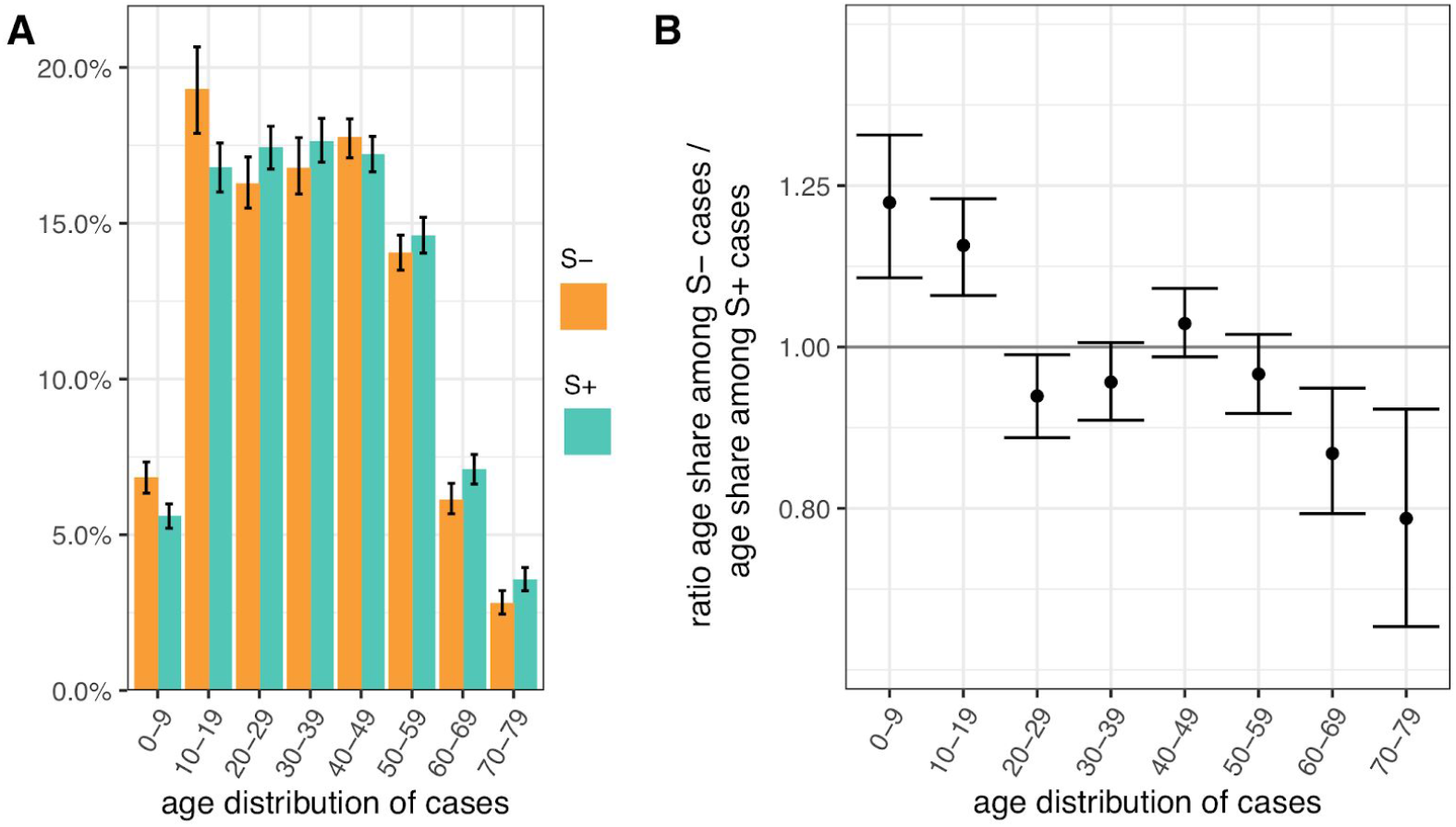
Age distribution of S-gene negative (S-) and S-gene positive (S+) PCR-positive pillar 2 cases from the SGSS dataset (not adjusted for TPR). Case numbers are weighted to compare S-cases from each NHS STP region and epidemiological week with an equal number of S+ cases from that STP and week (a case-control design), and standardised for differences in the age composition of each STP area. (A) Age distribution of S- and S+ cases. (B) Ratio of S- to S+ proportions of cases in each 10 year band. Results shown are for weeks 46-51. Ages were capped at 80. 95% empirical confidence intervals calculated by bootstrapping over STP areas and weeks, and sampling variation within STP areas and weeks.

### Regression analysis of VOC transmissibility

To investigate the effect of VOC frequency on the overall time-varying reproduction number, R_t_, we undertook a number of regression analyses. We conduct our analyses at two different spatial scales - lower tier local authority (LTLA) and NHS STP areas. For each, we estimated R_t_ by week and area using data on pillar 2 testing, deaths and hospitalisations using a previously described model^9,11^. Figure 5 shows the empirical relationship between weekly estimates of R_t_ at STP level and the frequency of the VOC estimates using genomic data.

**Figure 5:**
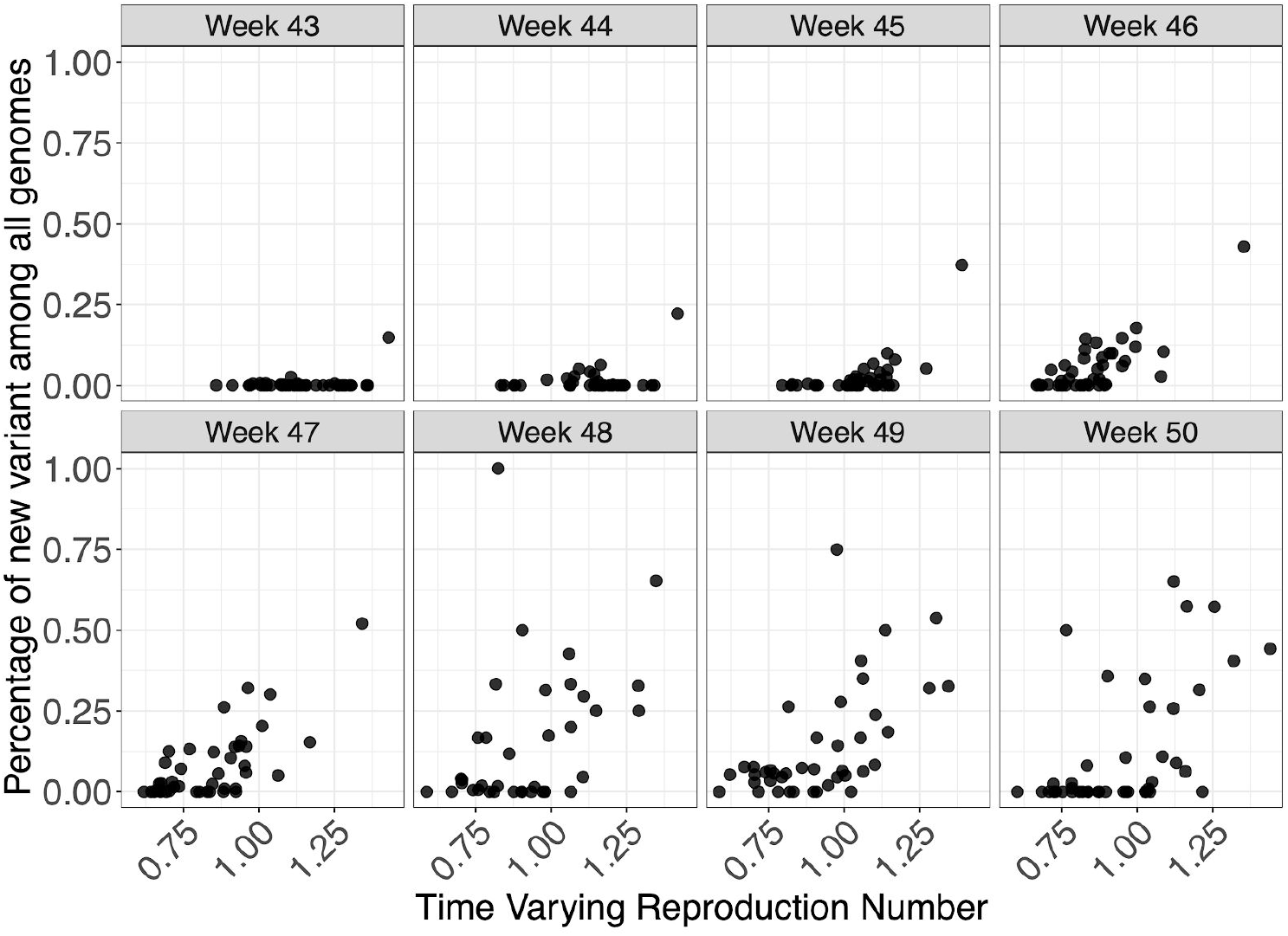
Relationship between genomic frequency of the VOC lineage among all genomes plotted against the time varying reproduction number for each week. Each datapoint is an STP area.

We apply a range of frequentist models with a bootstrapping procedure to account for non-normality in responses, as well as a Bayesian regression which explicitly models VOC frequency, such that it simultaneously informs the parameter for binomially-distributed observations of frequency and the R_t_ estimates. The role of geography in explaining variance of R_t_ was examined using both fixed and random effects. These models were applied to both genomic-based frequency estimates and TPR-adjusted SGTF proportions of pillar 2 cases for which S-gene data was available. Given this definition and the approximately 1 week generation time of SARS-CoV-2, we expect R_t_ to have stronger association with VOC frequency 1 week earlier. We therefore present regressions of R_t_ against frequency at week *t*-1 for our default analysis (where *t* spans weeks 44-50), and a regression of R_t_ against frequency at week *t* is provided in the Supplementary Information.

Regression results are reported in Table 2 (Table S2 for sensitivity analysis). We estimate the additive effect on R_t_, i.e., the increase or decrease in R_t_ (using R_t_ as response in the linear model) due to the variant. As an example, with an additive effect size of 0.4, an area with an R_t_ of 0.8 without the VOC would have an R_t_ of 1.2 if only the VOC was present. As expected, models which allow for fixed effects of week and region give lower effect sizes for the VOC than random effect models, given the latter constrain week and time effects more than fixed effect models, due to the assumptions that such effects arise from normal distributions. The Bayesian model results closely resemble those from the frequentist random effects model.

**Table 2.** Estimated additive change of reproduction numbers of VOC compared with other variants for different regression models, spatial resolutions, and data used to estimate the prevalence of the VOC. Analysis uses R_t_ estimates from weeks 44-50 and data on the proportion of the VOC one week earlier, to take account of the generation time of SARS-CoV-2.

| Model | Spatial Resolution | Data for Variants | Estimated effect [95% CI] |
| --- | --- | --- | --- |
| Fixed | STP | Genomic | 0.48 [0.31, 0.85] |
| Random | STP | Genomic | 0.67 [0.52, 1.11] |
| Bayes | STP | Genomic | 0.68 [0.44, 0.93] |
| Fixed | LTLA | TPR-adjusted SGTF | 0.42 [0.33, 0.58] |
| Random | LTLA | TPR-adjusted SGTF | 0.52 [0.45, 0.69] |
| Fixed | STP | TPR-adjusted SGTF | 0.36 [0.11, 0.58] |
| Random | STP | TPR-adjusted SGTF | 0.47 [0.25, 0.70] |
| Bayes | STP | TPR-adjusted SGTF | 0.48 [0.31, 0.63] |

The results in Table 2 show a clear association between the VOC and R_t_. However, this analysis cannot prove causality. The estimated additive effect is specific to the conditions that prevailed in England during the time period examined.

### Estimating reproduction numbers for VOC and non-VOC independently

We estimated the reproduction number of the VOC via phylodynamic analysis of whole genome sequences from Pillar 2 national SARS-CoV-2 testing, sampled up to December 6, 2020. First, we fitted a non-parametric *skygrowth* model ^12^ by maximum likelihood to 776 genomes that we selected from England in inverse proportion to the number of diagnosed cases sequenced in each region by week (see Supporting Methods). This model indicates that the effective population size of VOC 202012/01 grew at a relatively stable rate of 58% per week from September 20 to December 6, corresponding to a reproduction number of 1.59. Estimates of growth rate were insensitive to uncertainty in the molecular clock rate of evolution. Second, we fitted the model to genomes from four regions with more than fifty sequences, Kent (n=701), Greater London (n=606), Essex (n=131), and Norfolk (n=81). This regional analysis indicated growth rates ranging from 58% to 92% per week, corresponding to reproduction numbers between 1.56 and 1.95 (Figure S6). Finally, we carried out a Bayesian non-parametric coalescent analysis using the Skygrid model^13^ using the same set of 776 genomes. This analysis showed growth until the start of November followed by a plateau for the month of November coincident with the second English lockdown (Figure S7). This suggests the lockdown constrained growth of the VOC, but was insufficient to cause a reduction in incidence. To estimate parameter values we also estimated the initial growth rate of the VOC lineage under a parametric logistic growth coalescent model^14^. Under this model we estimated a growth rate of 71.5 per year, corresponding to a doubling time of 3.7 days (95% CrI: 2.4 – 4.9) and a reproduction number of 2.27 (1.84 – 2.73). By comparison, a simple exponential growth model over this entire period yields a growth rate of 27.9 with a doubling time of 9.1 days (7.4, 11.2) and reproductive number of 1.50 (1.40 – 1.60).

In a parallel epidemiological analysis, we estimated VOC and non-VOC pillar 2 case numbers by STP area using TPR-corrected SGTF frequencies applied to overall PHE pillar 2 case numbers. We then estimate R_t_ by week separately for VOC and non-VOC, using the same model previously used to generate overall (non lineage-stratified) R_t_ estimates^11^. We first fit the unstratified model to estimate the infection ascertainment ratio (numbers of infections being identified as positive cases) and infection seeding (initial infections in each region). For seeding, we use the estimated infections from our unstratified model. The mean number of daily infections for week 42 and 43 are used for seeding both VOC and non-VOC models. The fraction of SGTF cases is used to distribute infections for seeding between VOC and non-VOC in weeks 42 and 43. We then compute R_t_ estimates for weeks 45-50, to avoid the seeding assumptions affecting R_t_ estimates. Figure 6A shows the mean posterior difference between R_t_ estimates for VOC and non-VOC for week 48 and 50, while figure 6B shows plots median R_t_ estimates for VOC and non-VOC across all NHS regions for weeks 45-50. The R_t_ estimates for VOC are greater than those for non-VOC for 94% of STP-week pairs (points above the diagonal in Figure 6B). Figure S4 shows the mean posterior difference between R_t_ estimates for VOC and non-VOC for all weeks 45-50, while Figure S5 shows the ratio of R_t_ estimates. The mean R_t_ difference across weeks 45-50 is 0.51 [95% CrI: −0.09 − 1.10] which was computed from the set of 42×6 (STP x week) posteriors of R_t_ estimated for the VOC and non-VOC. The mean ratio of the estimated R_t_ for the VOC and non-VOC was 1.56 [95%CI: 0.92 − 2.28] for the same period, see Figure S5. Aggregating across all STPs we find that the mean R_t_ during the second English lockdown across all STPs was 1.45 [0.91-1.89] for the VOC and 0.92 [0.86-1.06] for non-VOC strains.

**Figure 6:**
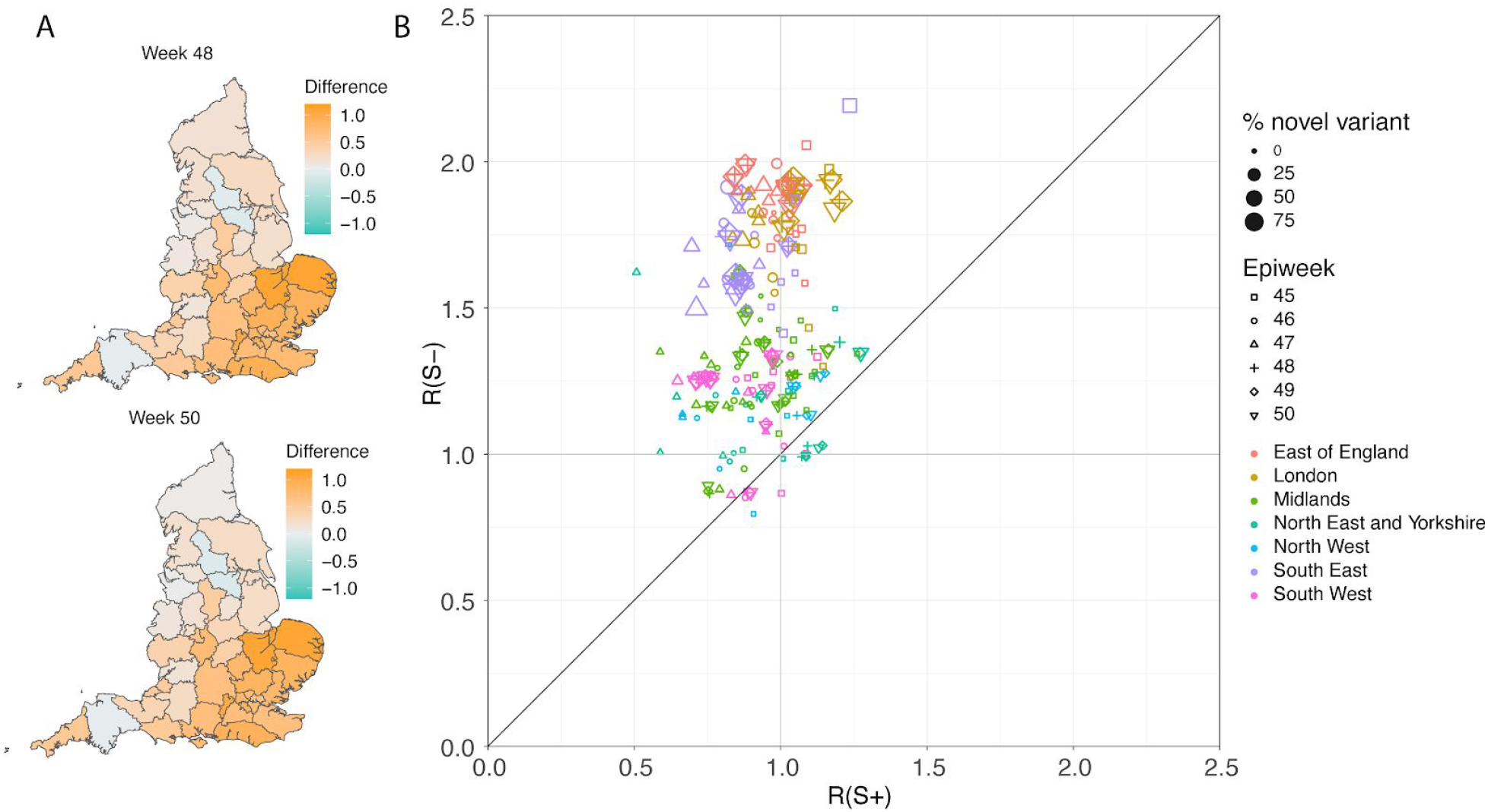
(A) Map of the difference in median R_t_ estimates for VOC and non-VOC variants for all STPs for weeks 48 and week 50. (B) Scatterplot of the reproduction numbers of VOC (S-) and non-VOC (S+) by STP and week. Point size indicates frequency of the VOC, while shape and colour signify week and NHS region, respectively.

## Discussion

While evidence has accumulated that substitutions associated with the B.1.1.7 lineage are associated with significant changes in virus phenotype^2–4,15^, assessing the extent to which these changes lead to meaningful differences in transmission between humans is challenging and cannot be evaluated experimentally. When randomised experimental studies are not possible, observational studies provide stronger evidence if consistent patterns are seen in multiple locations and at multiple times. While rapidly increasing frequency of a new lineage within a viral population is consistent with a selective advantage, it is also possible that increases in frequency may be caused by founder effects or genetic drift, especially for genetic variants which are repeatedly introduced from overseas^16,17^. But in contrast to previous genetic variants which have achieved high prevalence, we see expansion of the VOC from within the United Kingdom and a pattern of faster epidemic growth in tandem with expansion of the VOC has been repeated in multiple regions. In this paper we have focussed on spatiotemporally stratified analyses using a variety of statistical approaches to evaluate the relationship between SARS-CoV-2 transmission intensity and the frequency of the VOC, B.1.1.7 during November-December 2020 in different UK regions.

Assessment of the transmission characteristics of the VOC (B.1.1.7) was aided by the high correlation between its frequency and the occurrence of S-gene target failure (SGTF) in routine PCR testing of community cases of COVID-19 associated with the Δ69-70 deletion present in the VOC lineage (Figure 1 and S1). S-gene positivity results were available for over a third of all PCR-positive community COVID-19 cases for November and December 2020, allowing us to use SGTF frequency as a proxy for VOC frequency, and thus estimate VOC and non-VOC incidence trends by region over that time period. We see a very clear visual association between SGTF frequency and epidemic growth in nearly all areas (Figures 2 and S3), which is reinforced by empirical assessment of area-specific week on week growth factors of VOC and non-VOC case numbers (Figure 3) and by formal regression analyses of the association between estimates of local R_t_ and VOC frequency estimated from SGTF data (Table 2).

Finally, we used the SGTF data to independently estimate R_t_ by region and week for the VOC and non-VOC variants (Figures 6 and S4) and derived similar estimates for the increase in R_t_ associated with the VOC. This latter analysis is perhaps the most powerful, as no parametric assumptions are made about the relationship between R_t_ of the VOC and that of non-VOC strains.

Phylodynamic modelling provides additional information about growth of the VOC in October during a period when SGTF data is sparse. Although not apparent in all analyses, this suggests that the VOC expanded rapidly in October, with growth slowing (but not reversing) during national lockdown in November (Figures S6 and S7).

We were also able to rule out the hypothesis that increased incidence growth rates in the VOC are solely due to a change in the latent period or generation time distribution, but not the reproduction number itself (Table 1), since we see a large and statistically significant imbalance between regions where the VOC increased and where the non-VOC decreased, and vice-versa. A change solely in, for instance, the latent period would not be expected to change the direction of incidence growth.

We quantified the transmission advantage of the VOC relative to non-VOC lineages in two ways: as an additive increase in R that ranged between 0.4 and 0.7, and alternatively as a multiplicative increase in R that ranged between a 50% and 75% advantage. We were not able to distinguish between these two approaches in goodness-of-fit, and either is plausible mechanistically. A multiplicative transmission advantage would be expected if transmissibility had increased in all settings and individuals, while an additive advantage might reflect increases in transmissibility in specific subpopulations or contexts. More generally, the temporal context is important; these estimates of transmission advantage apply to a period where high levels of social distancing were in place in England; extrapolation to other transmission contexts, without detailed knowledge of the drivers of transmission, requires caution.

We observe a small but statistically significant shift towards under 20s being more affected by the VOC than non-VOC variants (Figure 4), even after controlling for variation by week and region. However, as with our earlier results, this observation does not resolve the mechanism that might underlie these differences. Differences between the age-distributions of VOC and non-VOC community cases may result from the overall increase in transmissibility of the VOC (especially during a time where lockdown was in force but schools were open), increased susceptibility of under 20s, or more apparent symptoms (and thus a propensity to seek testing) for the VOC in that age range.

There are a number of limitations to our analysis. The genomic and epidemiological data analysed was collected as part of routine surveillance, and thus may not be an entirely representative sample of SARS-CoV-2 infections in England over the time period considered. We also focussed on relatively simple, data-driven analyses using relatively simple models making parsimonious assumptions, rather than, for instance, attempting to model the long-term transmission dynamics of VOC and non-VOC lineages more mechanistically. We also did not attempt to explicitly model the spatiotemporal correlation intrinsic in infectious disease data, especially when considering the spread of a new variant from a point source. Doing so is an important priority for future work, but will require explicit incorporation of data on population movement patterns.

Early versions of our analyses informed the UK government policy response to this VOC and that of other countries. The substantial transmission advantage we have estimated the VOC to have over prior viral lineages poses major challenges for ongoing control of COVID-19 in the UK and elsewhere in the coming months. Social distancing measures will need to be more stringent than they would have otherwise. A particular concern is whether it will be possible to maintain control over transmission while allowing schools to reopen in January 2021. These policy questions will be informed by the ongoing urgent epidemiological investigation into this variant, most notably examining evidence for any changes in severity, but also giving more nuanced understanding into transmissibility changes, for instance in the household setting.

## Data Availability

All aggregated data to reproduce analysis will be provided in the url below.

https://github.com/mrc-ide/covid19-variant-N501Y

## Supplementary Information

### Methods

All code and data is available at https://github.com/mrc-ide/covid19-variant-N501Y

### Logistic growth model applied to variant frequency data

The logistic growth model fitted to VOC frequency data arises from a simple mechanistic model for competition between two strains. Let established lineages have a reproduction number *R* and let the VOC have reproduction number *R*(1+*s*). According to this model, the log odds of observing a variant over time will be proportional to (*R*/*g*)*st*, where *R* is assumed to be constant over weeks 44-49, *g* is the generation time, *s* is a selection coefficient (assumed to be constant) and t is time. If these conditions are met, s can be interpreted as a multiplicative change in the reproduction number *or* as a change in the generation time or as some combination of these factors. From available data on times of sampling each variant, the compound parameter (*R*/*g*)*s* can be estimated. An approximate estimate of *s* is obtained by treating *R* =1 and generation time *g*=6.5 days as a constant for weeks 44-49. In the text, we refer to s as the change in growth *per generation*, and is comparable to multiplicative changes in *R*_t_ estimated using other methods.

### Phylodynamic analysis

Analysis was based on Pillar 2 whole genome sequences with known Upper Tier Local Authority (UTLA). Maximum likelihood non-parametric phylodynamic analysis was carried out by: 1) estimating a maximum likelihood phylogeny in IQtree^18^ (HKY model of sequence evolution); 2) We removed duplicated identical sequences and estimated a time-scaled phylogeny using treedater^19^ using a strict molecular clock. The molecular clock rate of evolution was constrained to 0.0005 − 0.0015 substitutions per site per year. Small branch lengths in the tree were collapsed and polytomies randomly resolved to produce 20 new variations on the dated ML tree. 3) The skygrowth model was then fitted to these dated trees with parameters 64 time steps and tau bounded between 0.0025-20. Growth rate estimates were translated to reproduction numbers using the method of Wallinga and Lipsitch^20^ in the *epitrix* R package and using serial intervals from Flaxman et al.^9^ The main text reports the median growth rate over time. A sensitivity analysis was carried out with the molecular clock rate fixed at 0.001 substitutions per site per year which yielded only marginally different estimates of the median growth rate.

Sequence sample weights were used to select samples for phylodynamic modelling. Weights, assigned to sequence samples according to their UTLA and their collection date, correspond to the number of confirmed cases represented by each sequence in a UTLA relative to other UTLAs on the same date. To smooth over sparsity, case and sample counts were summed over the fourteen days prior to the date. Confirmed cases were accessed from the ONS API (https://api.coronavirus.data.gov.uk). Code to compute sequence sample weights is available at https://github.com/robj411/sequencing_coverage.

Bayesian coalescent phylodynamic analysis was performed using BEAST v1.10.4^14^ employing the Skygrid non-parametric approach ^21^ and exponential and logistic growth parametric models. A Jukes-Cantor model of substitution and a strict molecular clock were assumed. MCMC chains were run for 100M states, removing 10M as ‘burn-in’ and then thinning to 9000 samples from the posterior. Parameters were summarised and demographic curves reconstructed using Tracer v1.7.2 ^22^.

### SGTF as a biomarker for the VOC and frequency of SGTF over time

Data on SGTF among pillar 2 tests was obtained from the 3 largest (“Lighthouse”) PCR testing laboratories and integrated into the PHE Second Generation Surveillance System (SGSS) database. We also obtained the total number of cases reported by Public Health England.

Application of SGTF as a diagnostic for the VOC provides a large advantage over genomic sequencing in terms of cost, speed, and the sample size of available test results. We extracted 275,571 S target positive (S+) and 96,070 S target negative (S-) test results collected between 1 October and 19 December, 2020 and examined the potential to use SGTF cases (S-) as a biomarker for the VOC lineage. While the tests are not a representative sample of infections over this time period, they are a representative sample of tests within a given region and week and thus provide information about the relative abundance of the VOC versus other variants over time and between regions.

Other lineages have been observed to carry Δ69-70 which is associated with SGTF and which would have a similar impact on the TaqPath assay. The diagnostic specificity of SGTF will therefore vary over time and space as it depends on the abundance of other lineages with Δ69-70. This deletion is mostly found in global lineage B.1.258 and is highly linked with the spike N439K variant^23^. Lineage B.1.258 has circulated in the UK since June 2020 where it is now widespread. However, the frequency of this variant has been relatively stable since October 2020.

In order to capture diagnostic uncertainty with SGTF, we fitted a spatio-temporal model of the frequency of the VOC relative to other variants carrying Δ69-70. We fitted a generalized additive model^24,25^ to counts of genomes classified ^26^ as belonging to the VOC lineage or genomes which do not belong to the VOC lineage but still carry Δ69-70. Counts were tabulated by LTLA and epi week. We used a cubic spline to model trends over time, and correlation between neighbouring LTLAs was modeled with a Gaussian Markov Random Field (GMRF). This model was used to predict the true positive rate (TPR) − the probability that a sample collected at a particular time and place belongs to the VOC lineage, given that it carries Δ69-70. Figure S1 shows how this prediction has changed over weeks spanning November 2020. From week 50 onwards, we assumed the TPR was 1 across England. The fitted model thus provided an estimate of the true positive rate (TPR) of using SGTF as a proxy for VOC frequency as a function of time and region in England (Figure S2).

### Regression analysis of VOC transmissibility

Using reported COVID-19 PCR-positive case counts and deaths, we estimated the time-varying reproduction numbers R_t_ for epidemiological weeks 44-50 in each LTLA and STP in England. These estimates were obtained from a previously developed^11^ Bayesian semi-mechanistic transmission model with a latent weekly random walk process that does not include underlying factors that drive transmission. The STP model is based on pillar 2 cases only, and includes hospital admissions as an additional input; the LTLA model is based on both pillar 1 and pillar 2 data. Further model information can be found in previous publications^9,27^. These R_t_ estimates refer to the reproduction number of the infections that gave rise to the infections at time *t*. Given this definition and the approximately 1 week generation time of SARS-CoV-2, we would expect R_t_ to be most closely associated with VOC frequency 1 week earlier. We therefore present regressions of R_t_ against frequency at week *t*-1 for our default analysis (where *t* spans weeks 44-50), but present results for regression of R_t_ against frequency at week *t* as a sensitivity analysis in Supplementary Information.

We consider two data sources when estimating the proportion of the VOC: Genomic-based frequency estimates and TPR-adjusted SGTF proportions of pillar 2 cases for which S-gene data was available. For the SGTF-based estimates, frequency estimates were available for all STPs and LTLAs for the weeks 43-49 (294 STP-weeks in total). When using genomic data, 7 STP-weeks in the week range 43-49 had no sampled genomes, leaving 287 STP-weeks for analysis, for which we had an average of 137 genomes per STP week (of which on average 7 were the VOC).

We use two types of frequentist models. The first uses a fixed effect for each area, the second uses a random effect for each area. Fixed effects of epidemiological week were included in both cases. Confidence intervals for the fixed and random effects models were computed through a bootstrapping method which resamples the areas with replacement and also samples the counts of VOC in each week/area pair based on the observed counts. This bootstrap aims to account for potential skewness and non-normality of responses, dependence within areas and the randomness in the proportion of the VOC sampled within an area. When using genomic-based frequency estimates we excluded STP-weeks that had fewer than 5 genetic profiles, as these were deemed to not give reliable estimates of the proportion of VOC. This removed a further 14 out of the 287 STP-weeks from those analyses.

We also implemented a Bayesian regression model ^28^ at STP level, taking into account uncertainty in the frequency estimates and uncertainty in the R_t_ estimates. VOC frequency was modelled explicitly, such that it simultaneously informed the parameter for binomially-distributed observations of frequency and the R_t_ estimates. The regression for R_t_ included terms for the VOC frequency, the week (modelled with a smooth spline) and the area as a factor. The formula to describe the binomial frequency data includes a linear function of time with an area-dependent slope, and was fitted to the genome counts and the counts of S-negative cases separately.

**Table S1.** Estimated growth difference per generation of B.1.1.7 by English administrative region based on a maximum likelihood estimation of logistic growth in variant frequency and assuming a 6.5 day generation time and R_t_=1 for established lineages. Confidence intervals are based on likelihood profiles

|  | MLE %Growth difference<br>per generation (95%<br>confidence interval) | SGTF MLE<br>%Growth difference<br>per generation |
| --- | --- | --- |
| South East | 49 (43-55) | 54 |
| London | 49 (43-54) | 60 |
| East of England | 53 (44-62) | 67 |
| West Midlands |  | 59 |
| East Midlands |  | 56 |
| North West |  | 58 |
| Yorkshire and<br>The Humber |  | 40 |
| North East |  | 75 |
| South West |  | 48 |

**Table S2.** Estimated additive change of reproduction numbers of VOC compared with non-VOC using different regression models, spatial resolutions, and data to estimate the prevalence of the VOC. Analyses use data on the proportion of the VOC and estimates of R_t_ in weeks 44-50.

| Model | Spatial Resolution | Data for Variants | Estimated effect [95% CI] |
| --- | --- | --- | --- |
| Fixed | STP | Genomic | 0.34 [0.16, 0.60] |
| Random | STP | Genomic | 0.52 [0.35, 0.85] |
| Bayes | STP | Genomic | 0.54 [0.33, 0.77] |
| Fixed | LTLA | TPR-adjusted SGTF | 0.45 [0.40, 0.59] |
| Random | LTLA | TPR-adjusted SGTF | 0.53 [0.49, 0.67] |
| Fixed | STP | TPR-adjusted SGTF | 0.35 [0.15, 0.56] |
| Random | STP | TPR-adjusted SGTF | 0.44 [0.27, 0.65] |
| Bayes | STP | TPR-adjusted SGTF | 0.43 [0.30, 0.56] |

**Figure S1.**
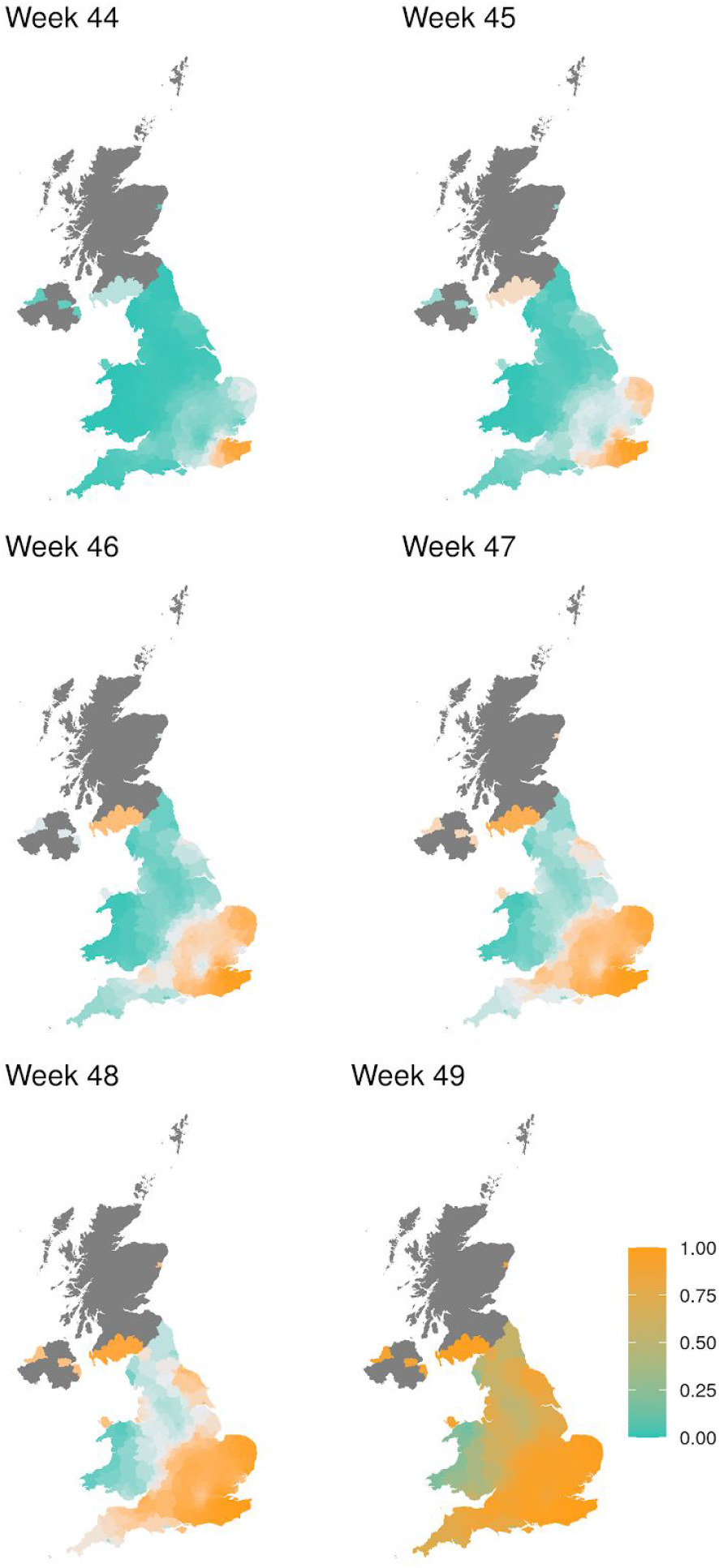
Estimate of true positive rates for classification of B.1.1.7 infection given SGTF result (S-) as a function of time and UK region. The colour gradient shows the probability of sampling a B.1.1.7 sequence conditional on sampling any sequence with Δ69-70.

**Figure S2.**
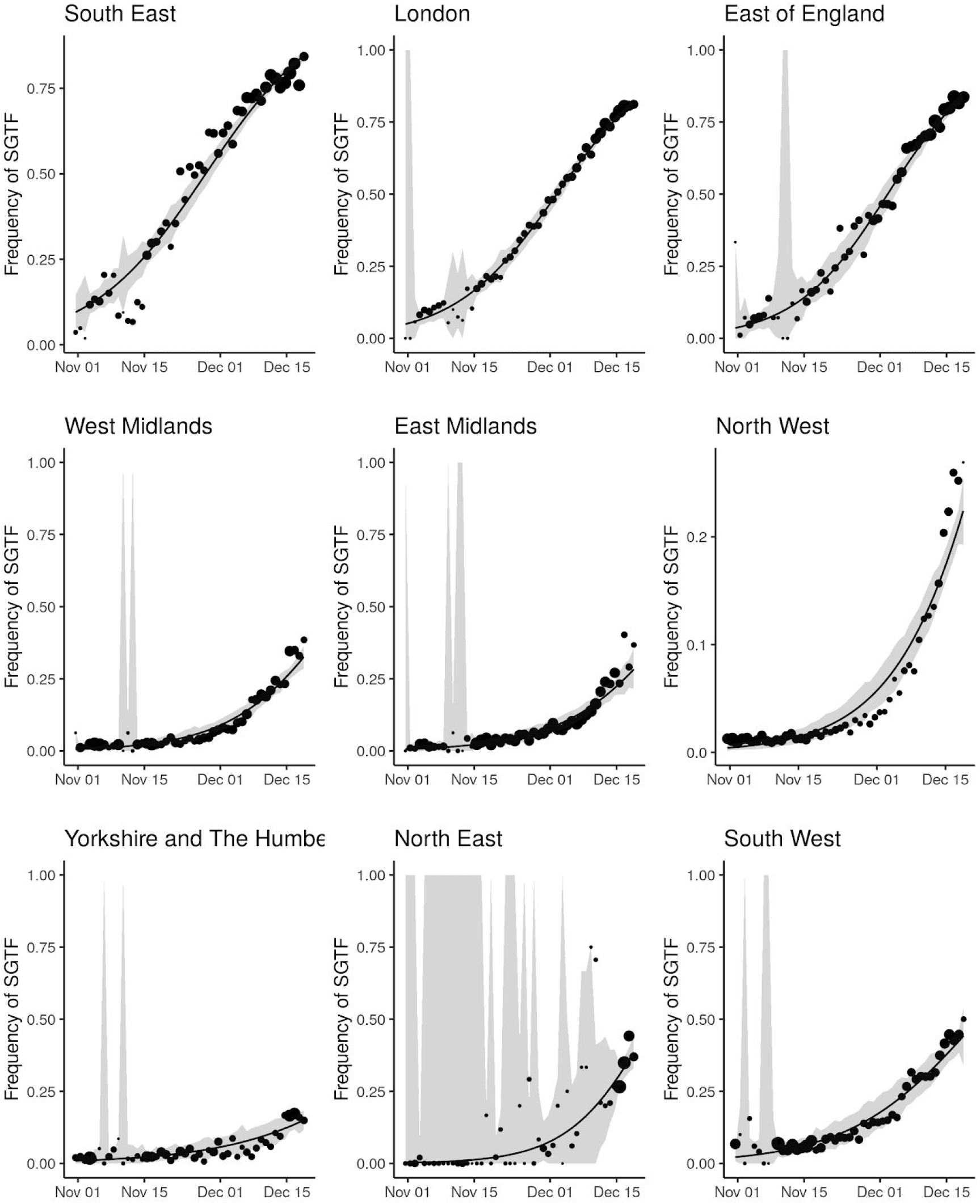
Empirical (point) and estimated (line) frequencies of TPR-adjusted SGTF frequencies over time. The size of points correspond to the number of samples observed by day. Confidence intervals show 95% estimated sampling error for daily proportions.

**Figure S3.**
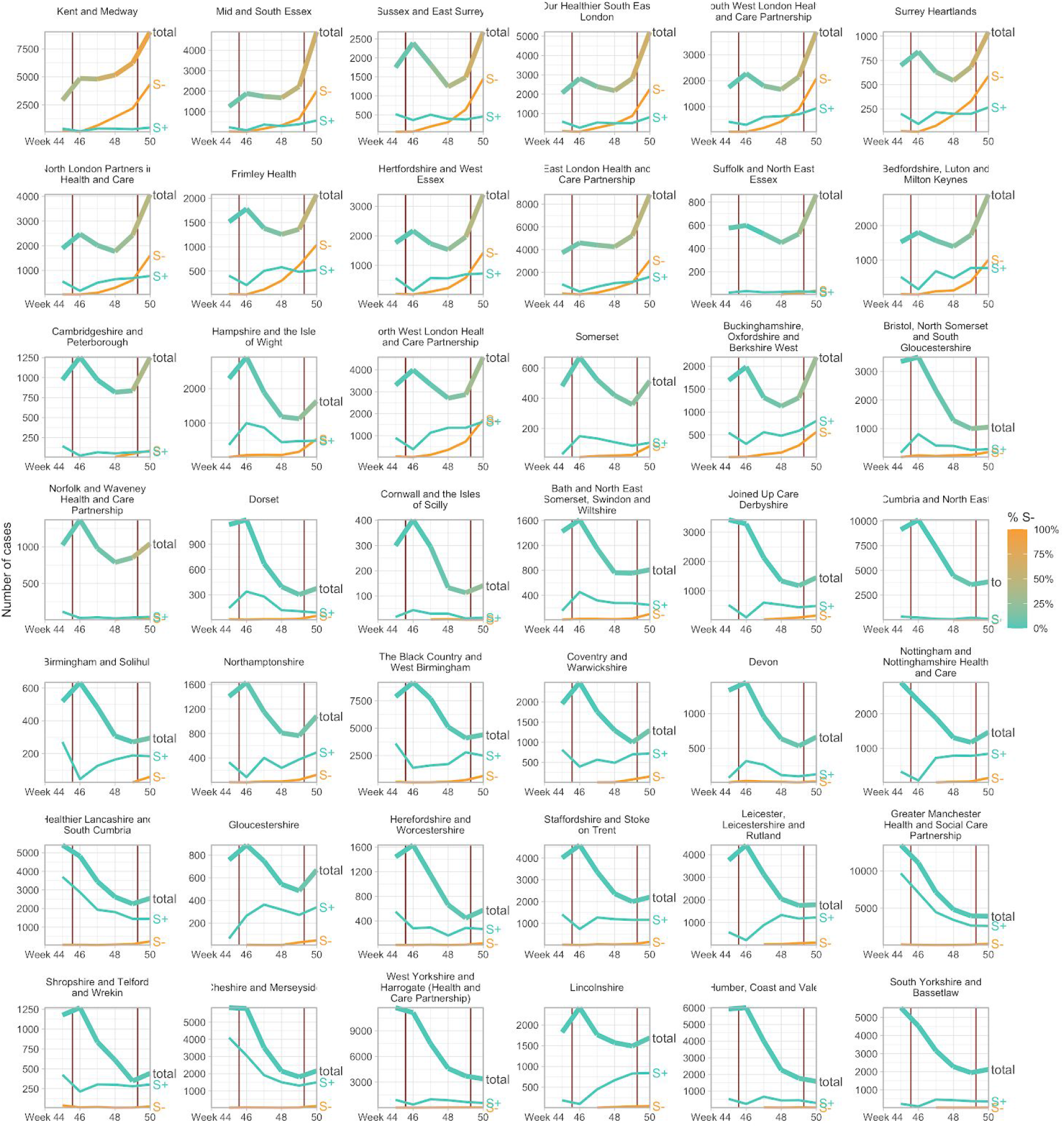
Case trends in all NHS STP areas, ordered by decreasing frequency of S-in the last week shown. Total cases reported are shown as a thick line. A subset of these - those tested in the 3 largest “Lighthouse” laboratories - were tested for SGTF. The total cases line is coloured according to percentage S-among those tested. Counts of S+ and S-reported via the PHE SGSS system are shown by the thin lines. The dates of the second lockdown are indicated by the vertical red lines. Raw SGTF data are shown here (not adjusted for TPR), so S-cases in earlier weeks include other non-VOC lineages, especially outside the East and South East of England.

**Figure S4:**
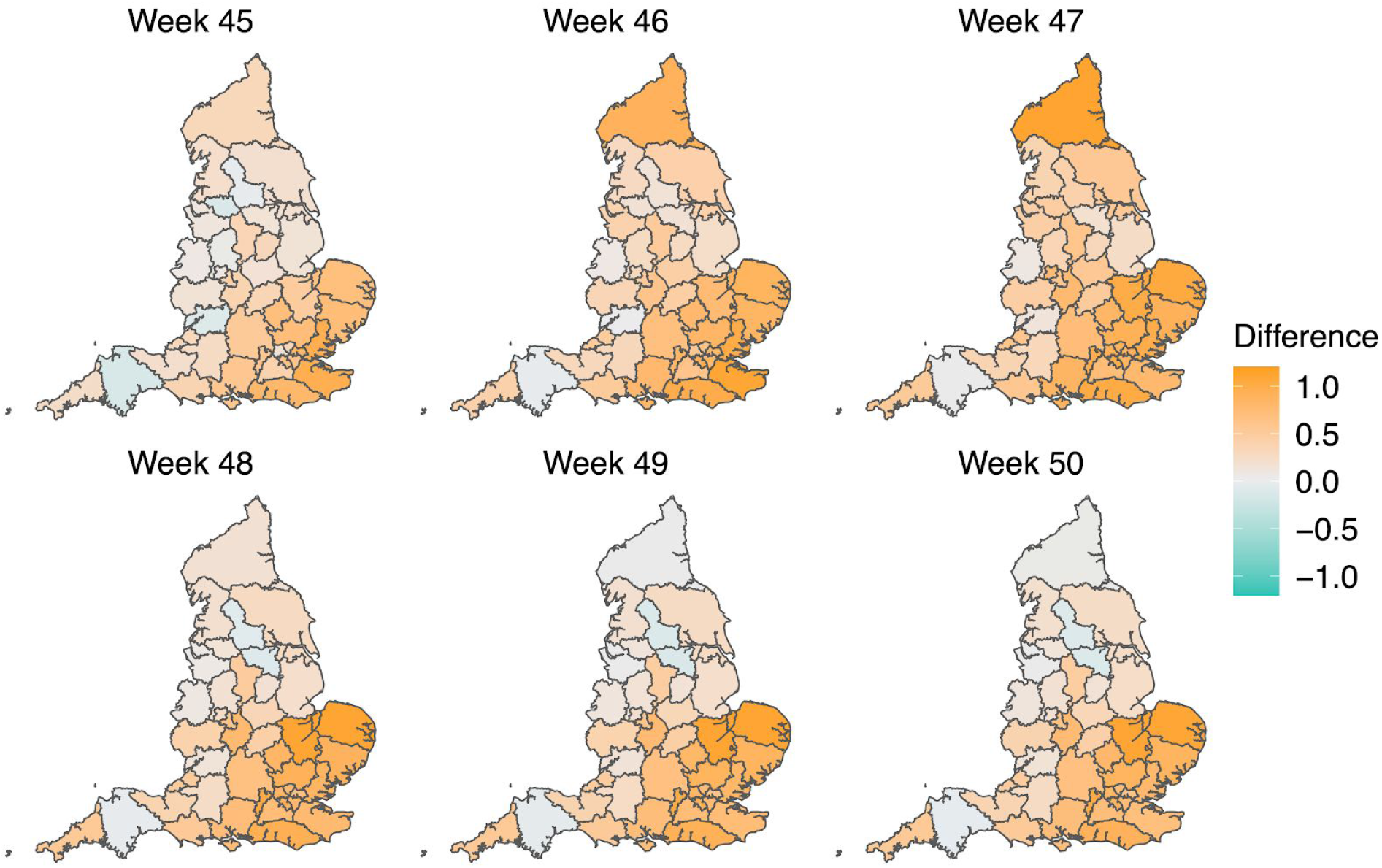
Map of the difference in median Rt estimates for VOC and non-VOC variants for all STPs between week 45 to week 50. The darker orange color indicates the additive advantage VOC has over non-VOC variant for Rt, whereas the darker green color shows the advantage for non-VOC variant over VOC.

**Figure S5.**
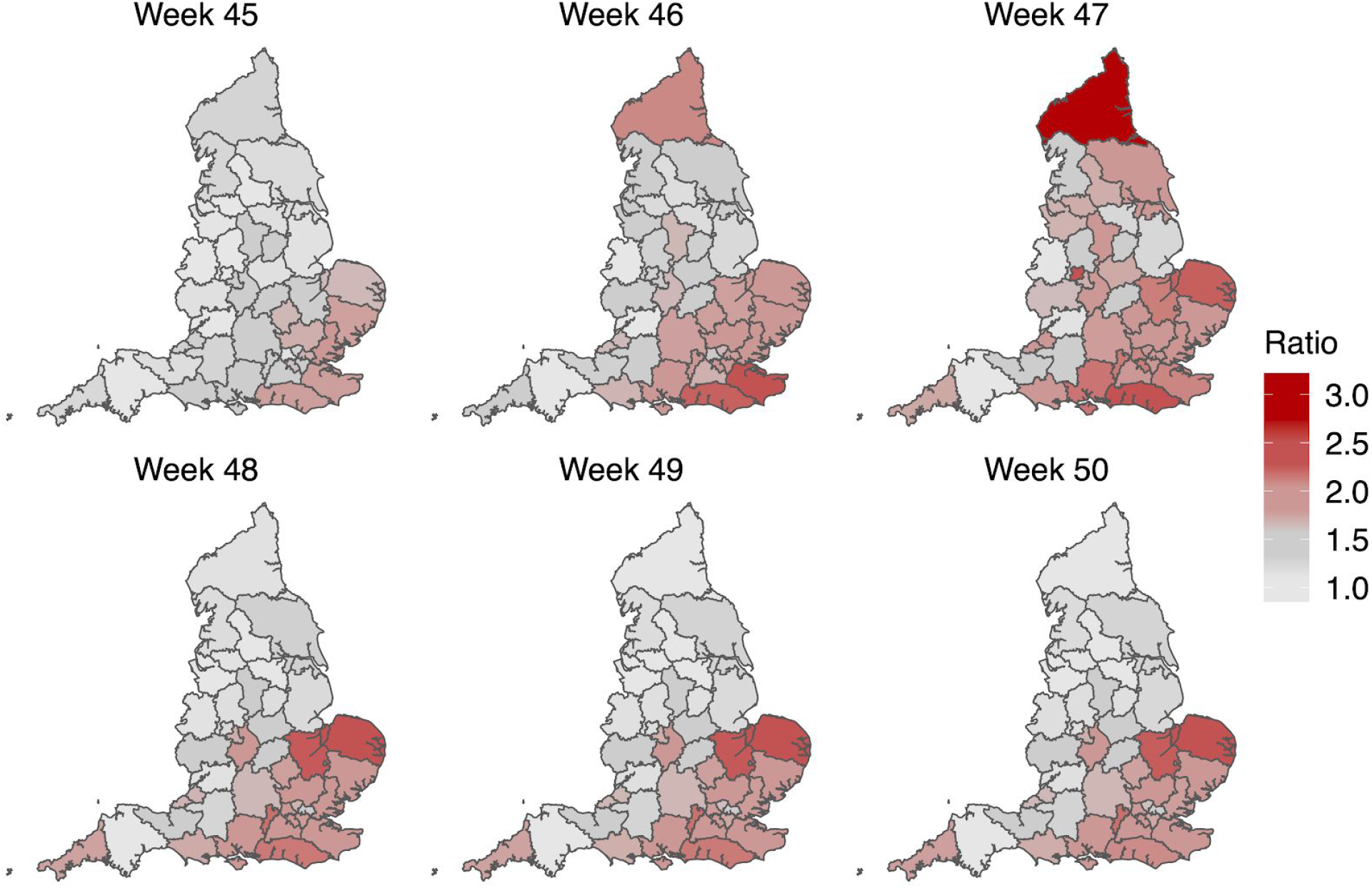
Map of the ratio of median Rt estimates for VOC and non-VOC variants for all STPs between week 45 to week 50. Darker color indicates the higher multiplicative advantage for VOC variant in comparison to the non-VOC variant. The mean of the ratio between R estimates for S- and S+ for all posterior samples across weeks 45 to 50 and all STPs is 1.56, with 95% CI [0.92 − 2.28]^1^.

**Figure S6.**
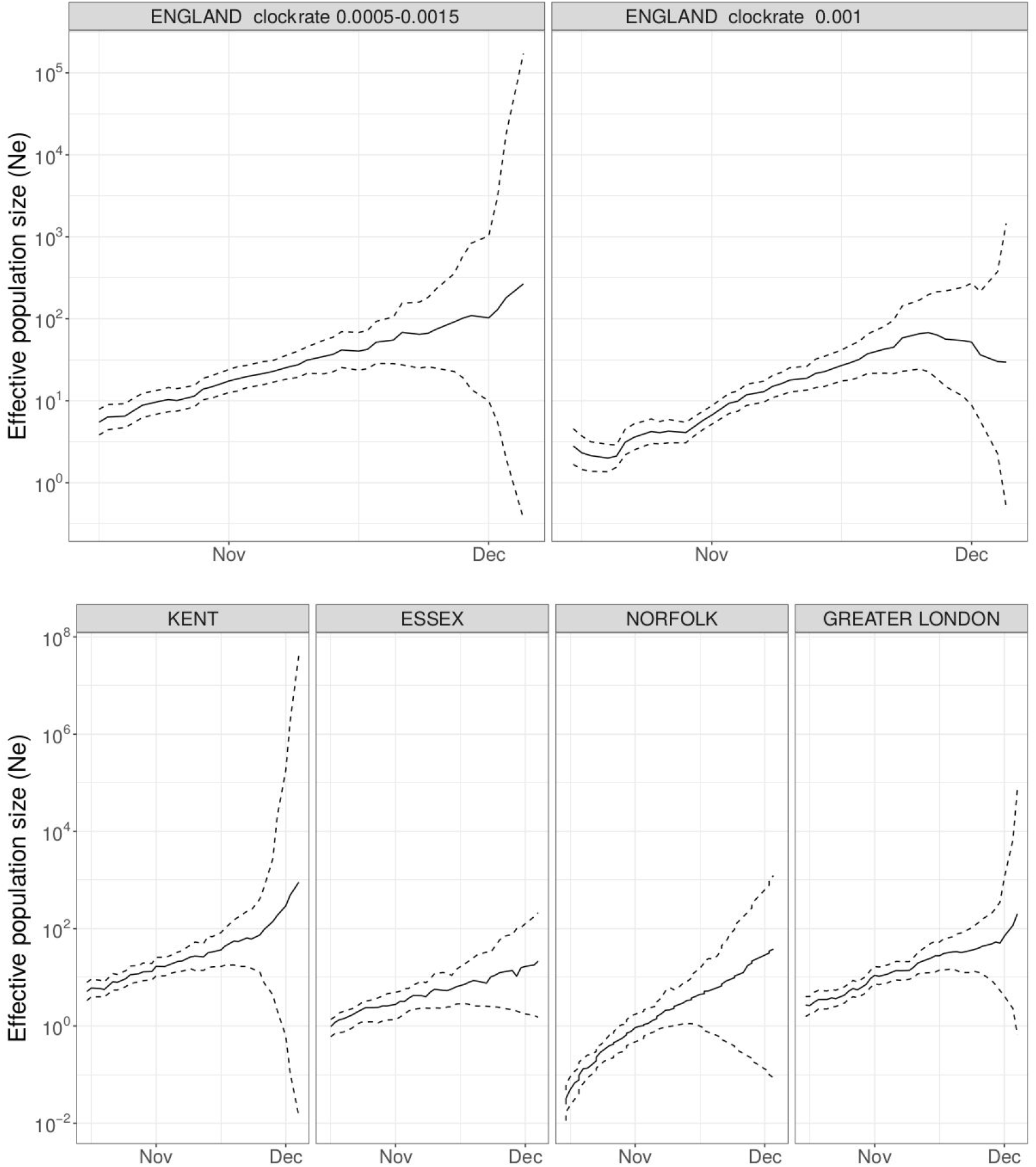
Maximum likelihood *skygrowth* estimates of the effective population size of the VOC through time in England and in four areas of England with more than 80 whole genome sequences. In the upper right panel the molecular clock rate of evolution was fixed at 0.001 substitutions per site per year. In all other analyses the rate was estimated and bound between 0.0005 and 0.0015 substitutions per site per year.

**Figure S7.**
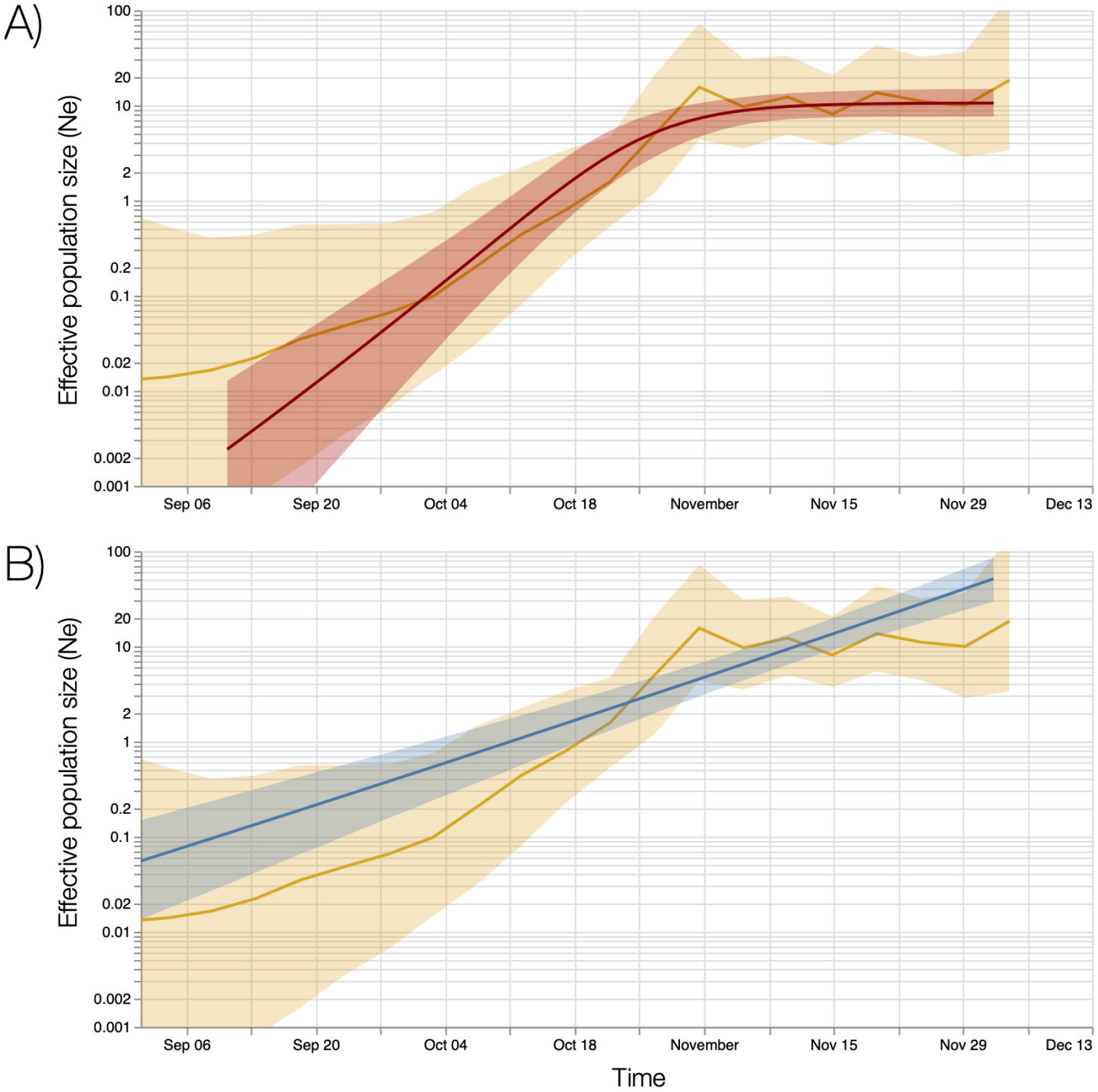
Bayesian estimates of effective population size through time based on 776 genomes sampled between October and December 6. Top and bottom panels show estimates based on a Bayesian skygrid model overlaid with fits of different parametric models. A) Estimates based on a logistic growth model. B) Estimates from an exponential growth model.

## Supplementary Appendix: COG-UK consortium members

**Funding acquisition, Leadership and supervision, Metadata curation, Project administration, Samples and logistics, Sequencing and analysis, Software and analysis tools, and Visualisation:** Dr Samuel C Robson ^13^.

**Funding acquisition, Leadership and supervision, Metadata curation, Project administration, Samples and logistics, Sequencing and analysis, and Software and analysis tools:**

Prof Nicholas J Loman ^41^, Dr Thomas R Connor ^10, 69^.

**Leadership and supervision, Metadata curation, Project administration, Samples and logistics, Sequencing and analysis, Software and analysis tools, and Visualisation:**

Dr Tanya Golubchik ^5^.

**Funding acquisition, Metadata curation, Samples and logistics, Sequencing and analysis, Software and analysis tools, and Visualisation:**

Dr Rocio T Martinez Nunez ^42^.

**Funding acquisition, Leadership and supervision, Metadata curation, Project administration, and Samples and logistics:**

Dr Catherine Ludden ^88^.

**Funding acquisition, Leadership and supervision, Metadata curation, Samples and logistics, and Sequencing and analysis:**

Dr Sally Corden ^69^.

**Funding acquisition, Leadership and supervision, Project administration, Samples and logistics, and Sequencing and analysis:**

Ian Johnston ^99^ and Dr David Bonsall ^5^.

**Funding acquisition, Leadership and supervision, Sequencing and analysis, Software and analysis tools, and Visualisation:**

Prof Colin P Smith ^87^ and Dr Ali R Awan ^28^.

**Funding acquisition, Samples and logistics, Sequencing and analysis, Software and analysis tools, and Visualisation:**

Dr Giselda Bucca ^87^.

**Leadership and supervision, Metadata curation, Project administration, Samples and logistics, and Sequencing and analysis:**

Dr M. Estee Torok ^22, 101^.

**Leadership and supervision, Metadata curation, Project administration, Samples and logistics, and Visualisation:**

Dr Kordo Saeed ^81, 110^ and Dr Jacqui A Prieto ^83, 109^.

**Leadership and supervision, Metadata curation, Project administration, Sequencing and analysis, and Software and analysis tools:**

Dr David K Jackson ^99^.

**Metadata curation, Project administration, Samples and logistics, Sequencing and analysis, and Software and analysis tools:**

Dr William L Hamilton ^22^.

**Metadata curation, Project administration, Samples and logistics, Sequencing and analysis, and Visualisation:**

Dr Luke B Snell ^11^.

**Funding acquisition, Leadership and supervision, Metadata curation, and Samples and logistics:**

Dr Catherine Moore ^69^.

**Funding acquisition, Leadership and supervision, Project administration**, and **Samples and logistics:**

Dr Ewan M Harrison ^99, 88^.

**Leadership and supervision, Metadata curation, Project administration, and Samples and logistics:**

Dr Sonia Goncalves ^99^.

**Leadership and supervision, Metadata curation, Samples and logistics, and Sequencing and analysis:**

Prof Ian G Goodfellow ^24^, Dr Derek J Fairley ^3, 72^, Prof Matthew W Loose ^18^ and Joanne Watkins ^69^.

**Leadership and supervision, Metadata curation, Samples and logistics, and Software and analysis tools:**

Rich Livett ^99^.

**Leadership and supervision, Metadata curation, Samples and logistics, and Visualisation:**

Dr Samuel Moses ^25, 106^.

**Leadership and supervision, Metadata curation, Sequencing and analysis, and Software and analysis tools:**

Dr Roberto Amato ^99^, Dr Sam Nicholls ^41^ and Dr Matthew Bull ^69^.

**Leadership and supervision, Project administration, Samples and logistics, and Sequencing and analysis:**

Prof Darren L Smith ^37, 58, 105^.

**Leadership and supervision, Sequencing and analysis, Software and analysis tools, and Visualisation:**

Dr Jeff Barrett ^99^ and Prof David M Aanensen ^14, 114^.

**Metadata curation, Project administration, Samples and logistics, and Sequencing and analysis:**

Dr Martin D Curran ^65^, Dr Surendra Parmar ^65^, Dr Dinesh Aggarwal ^95, 99, 64^ and Dr James G Shepherd ^48^.

**Metadata curation, Project administration, Sequencing and analysis, and Software and analysis tools:**

Dr Matthew D Parker ^93^.

**Metadata curation, Samples and logistics, Sequencing and analysis, and Visualisation:**

Dr Sharon Glaysher ^61^.

**Metadata curation, Sequencing and analysis, Software and analysis tools, and Visualisation:** Dr Matthew Bashton ^37, 58^, Dr Anthony P Underwood ^14, 114^, Dr Nicole Pacchiarini ^69^ and Dr Katie F Loveson ^77^.

**Project administration, Sequencing and analysis, Software and analysis tools, and Visualisation:**

Dr Alessandro M Carabelli ^88^.

**Funding acquisition, Leadership and supervision, and Metadata curation:**

Dr Kate E Templeton ^53, 90^.

**Funding acquisition, Leadership and supervision, and Project administration:**

Dr Cordelia F Langford ^99^, John Sillitoe ^99^, Dr Thushan I de Silva ^93^ and Dr Dennis Wang ^93^.

**Funding acquisition, Leadership and supervision, and Sequencing and analysis:**

Prof Dominic Kwiatkowski ^99, 107^, Prof Andrew Rambaut ^90^, Dr Justin O’Grady ^70, 89^ and Dr Simon Cottrell ^69^.

**Leadership and supervision, Metadata curation, and Sequencing and analysis:**

Prof Matthew T.G. Holden ^68^ and Prof Emma C Thomson ^48^.

**Leadership and supervision, Project administration, and Samples and logistics:**

Dr Husam Osman ^64, 36^, Dr Monique Andersson ^59^, Prof Anoop J Chauhan ^61^ and Dr Mohammed O Hassan-Ibrahim ^6^.

**Leadership and supervision, Project administration, and Sequencing and analysis:**

Dr Mara Lawniczak ^99^.

**Leadership and supervision, Samples and logistics, and Sequencing and analysis:**

Prof Ravi Kumar Gupta ^88, 113^, Dr Alex Alderton ^99^, Dr Meera Chand ^66^, Dr Chrystala Constantinidou ^94^, Dr Meera Unnikrishnan ^94^, Prof Alistair C Darby ^92^, Prof Julian A Hiscox ^92^ and Prof Steve Paterson ^92^.

**Leadership and supervision, Sequencing and analysis, and Software and analysis tools:**

Dr Inigo Martincorena ^99^, Prof David L Robertson ^48^, Dr Erik M Volz ^39^, Dr Andrew J Page ^70^ and Prof Oliver G Pybus ^23^.

**Leadership and supervision, Sequencing and analysis, and Visualisation:**

Dr Andrew R Bassett ^99^.

**Metadata curation, Project administration, and Samples and logistics:**

Dr Cristina V Ariani ^99^, Dr Michael H Spencer Chapman ^99, 88^, Dr Kathy K Li ^48^, Dr Rajiv N Shah ^48^, Dr Natasha G Jesudason ^48^ and Dr Yusri Taha ^50^.

**Metadata curation, Project administration, and Sequencing and analysis:**

Martin P McHugh ^53^ and Dr Rebecca Dewar ^53^.

**Metadata curation, Samples and logistics, and Sequencing and analysis:**

Dr Aminu S Jahun ^24^, Dr Claire McMurray ^41^, Ms Sarojini Pandey ^84^, Dr James P McKenna ^3^, Dr Andrew Nelson ^58, 105^, Dr Gregory R Young ^37, 58^, Dr Clare M McCann ^58, 105^ and Mr Scott Elliott ^61^.

**Metadata curation, Samples and logistics, and Visualisation:**

Ms Hannah Lowe ^25^.

**Metadata curation, Sequencing and analysis, and Software and analysis tools:**

Dr Ben Temperton ^91^, Dr Sunando Roy ^82^, Dr Anna Price ^10^, Dr Sara Rey ^69^ and Mr Matthew Wyles ^93^.

**Metadata curation, Sequencing and analysis, and Visualisation:**

Stefan Rooke ^90^ and Dr Sharif Shaaban ^68^.

**Project administration, Samples and logistics, Sequencing and analysis:**

Dr Mariateresa de Cesare ^98^.

**Project administration, Samples and logistics, and Software and analysis tools:**

Laura Letchford ^99^.

**Project administration, Samples and logistics, and Visualisation:**

Miss Siona Silveira ^81^, Dr Emanuela Pelosi ^81^ and Dr Eleri Wilson-Davies ^81^.

**Samples and logistics, Sequencing and analysis, and Software and analysis tools:**

Dr Myra Hosmillo ^24^.

**Sequencing and analysis, Software and analysis tools, and Visualisation:**

Áine O’Toole ^90^, Dr Andrew R Hesketh ^87^, Mr Richard Stark ^94^, Dr Louis du Plessis ^23^, Dr Chris Ruis ^88^, Dr Helen Adams ^4^ and Dr Yann Bourgeois ^76^.

**Funding acquisition, and Leadership and supervision:**

Dr Stephen L Michell ^91^, Prof Dimitris Gramatopoulos ^84, 112^, Dr Jonathan Edgeworth ^12^, Prof Judith Breuer

^30, 82^, Prof John A Todd ^98^ and Dr Christophe Fraser ^5^.

**Funding acquisition, and Project administration:**

Dr David Buck ^98^ and Michaela John ^9^.

**Leadership and supervision, and Metadata curation:**

Dr Gemma L Kay ^70^.

**Leadership and supervision, and Project administration:**

Steve Palmer ^99^, Prof Sharon J Peacock ^88, 64^ and David Heyburn ^69^.

**Leadership and supervision, and Samples and logistics:**

Danni Weldon ^99^, Dr Esther Robinson ^64, 36^, Prof Alan McNally ^41, 86^, Dr Peter Muir ^64^, Dr Ian B Vipond ^64^, Dr John BoYes ^29^, Dr Venkat Sivaprakasam ^46^, Dr Tranprit Saluja ^75^, Dr Samir Dervisevic ^54^ and Dr Emma J Meader ^54^.

**Leadership and supervision, and Sequencing and analysis:**

Dr Naomi R Park ^99^, Karen Oliver ^99^, Dr Aaron R Jeffries ^91^, Dr Sascha Ott ^94^, Dr Ana da Silva Filipe ^48^, Dr David A Simpson ^72^ and Dr Chris Williams ^69^.

**Leadership and supervision, and Visualisation:**

Dr Jane A H Masoli ^73, 91^.

**Metadata curation, and Samples and logistics:**

Dr Bridget A Knight ^73, 91^, Dr Christopher R Jones ^73, 91^, Mr Cherian Koshy ^1^, Miss Amy Ash ^1^, Dr Anna Casey ^71^, Dr Andrew Bosworth ^64, 36^, Dr Liz Ratcliffe ^71^, Dr Li Xu-McCrae ^36^, Miss Hannah M Pymont ^64^, Ms Stephanie Hutchings ^64^, Dr Lisa Berry ^84^, Ms Katie Jones ^84^, Dr Fenella Halstead ^46^, Mr Thomas Davis ^21^, Dr Christopher Holmes ^16^, Prof Miren Iturriza-Gomara ^92^, Dr Anita O Lucaci ^92^, Dr Paul Anthony Randell ^38, 104^, Dr Alison Cox ^38, 104^, Pinglawathee Madona ^38, 104^, Dr Kathryn Ann Harris ^30^, Dr Julianne Rose Brown ^30^, Dr Tabitha W Mahungu ^74^, Dr Dianne Irish-Tavares ^74^, Dr Tanzina Haque ^74^, Dr Jennifer Hart ^74^, Mr Eric Witele ^74^, Mrs Melisa Louise Fenton ^75^, Mr Steven Liggett ^79^, Dr Clive Graham ^56^, Ms Emma Swindells ^57^, Ms Jennifer Collins ^50^, Mr Gary Eltringham ^50^, Ms Sharon Campbell ^17^, Dr Patrick C McClure ^97^, Dr Gemma Clark ^15^, Dr Tim J Sloan ^60^, Mr Carl Jones ^15^ and Dr Jessica Lynch ^2, 111^.

**Metadata curation, and Sequencing and analysis:**

Dr Ben Warne ^8^, Steven Leonard ^99^, Jillian Durham ^99^, Dr Thomas Williams ^90^, Dr Sam T Haldenby ^92^, Dr Nathaniel Storey ^30^, Dr Nabil-Fareed Alikhan ^70^, Dr Nadine Holmes ^18^, Dr Christopher Moore ^18^, Mr Matthew Carlile ^18^, Malorie Perry ^69^, Dr Noel Craine ^69^, Prof Ronan A Lyons ^80^, Miss Angela H Beckett ^13^, Salman Goudarzi ^77^, Christopher Fearn ^77^, Kate Cook ^77^, Hannah Dent ^77^ and Hannah Paul ^77^.

**Metadata curation, and Software and analysis tools:**

Robert Davies ^99^.

**Project administration, and Samples and logistics:**

Beth Blane ^88^, Sophia T Girgis ^88^, Dr Mathew A Beale ^99^, Katherine L Bellis ^99, 88^, Matthew J Dorman ^99^, Eleanor Drury ^99^, Leanne Kane ^99^, Sally Kay ^99^, Dr Samantha McGuigan ^99^, Dr Rachel Nelson ^99^, Liam Prestwood ^99^, Dr Shavanthi Rajatileka ^99^, Dr Rahul Batra ^12^, Dr Rachel J Williams ^82^, Dr Mark Kristiansen ^82^, Dr Angie Green ^98^, Miss Anita Justice ^59^, Dr Adhyana I.K Mahanama ^81, 102^ and Dr Buddhini Samaraweera ^81, 102^.

**Project administration, and Sequencing and analysis:**

Dr Nazreen F Hadjirin ^88^ and Dr Joshua Quick ^41^.

**Project administration, and Software and analysis tools:**

Mr Radoslaw Poplawski ^41^.

**Samples and logistics, and Sequencing and analysis:**

Leanne M Kermack ^88^, Nicola Reynolds ^7^, Grant Hall ^24^, Yasmin Chaudhry ^24^, Malte L Pinckert ^24^, Dr Iliana Georgana ^24^, Dr Robin J Moll ^99^, Dr Alicia Thornton ^66^, Dr Richard Myers ^66^, Dr Joanne Stockton ^41^, Miss Charlotte A Williams ^82^, Dr Wen C Yew ^58^, Alexander J Trotter ^70^, Miss Amy Trebes ^98^, Mr George MacIntyre-Cockett ^98^, Alec Birchley ^69^, Alexander Adams ^69^, Amy Plimmer ^69^, Bree Gatica-Wilcox ^69^, Dr Caoimhe McKerr ^69^, Ember Hilvers ^69^, Hannah Jones ^69^, Dr Hibo Asad ^69^, Jason Coombes ^69^, Johnathan M Evans ^69^, Laia Fina ^69^, Lauren Gilbert ^69^, Lee Graham ^69^, Michelle Cronin ^69^, Sara

Kumziene-SummerhaYes ^69^, Sarah Taylor ^69^, Sophie Jones ^69^, Miss Danielle C Groves ^93^, Mrs Peijun Zhang ^93^, Miss Marta Gallis ^93^ and Miss Stavroula F Louka ^93^.

**Samples and logistics, and Software and analysis tools:**

Dr Igor Starinskij ^48^.

**Sequencing and analysis, and Software and analysis tools:**

Dr Chris J Illingworth ^47^, Dr Chris Jackson ^47^, Ms Marina Gourtovaia ^99^, Gerry Tonkin-Hill ^99^, Kevin Lewis ^99^, Dr Jaime M Tovar-Corona ^99^, Dr Keith James ^99^, Dr Laura Baxter ^94^, Dr Mohammad T. Alam ^94^, Dr Richard J Orton ^48^, Dr Joseph Hughes ^48^, Dr Sreenu Vattipally ^48^, Dr Manon Ragonnet-Cronin ^39^, Dr Fabricia F. Nascimento ^39^, Mr David Jorgensen ^39^, Ms Olivia Boyd ^39^, Ms Lily Geidelberg ^39^, Dr Alex E Zarebski ^23^, Dr Jayna Raghwani ^23^, Dr Moritz UG Kraemer ^23^, Joel Southgate ^10, 69^, Dr Benjamin B Lindsey ^93^ and Mr Timothy M Freeman ^93^.

**Software and analysis tools, and Visualisation:**

Jon-Paul Keatley ^99^, Dr Joshua B Singer ^48^, Leonardo de Oliveira Martins ^70^, Dr Corin A Yeats ^14^, Dr Khalil Abudahab ^14, 114^, Mr Ben EW Taylor ^14, 114^ and Mirko Menegazzo ^14^.

**Leadership and supervision:**

Prof John Danesh ^99^, Wendy Hogsden ^46^, Dr Sahar Eldirdiri ^21^, Mrs Anita Kenyon ^21^, Dr Jenifer Mason ^43^, Mr Trevor I Robinson ^43^, Prof Alison Holmes ^38, 103^, Dr James Price ^38, 103^, Prof John A Hartley ^82^, Dr Tanya Curran ^3^, Dr Alison E Mather ^70^, Dr Giri Shankar ^69^, Dr Rachel Jones ^69^, Dr Robin Howe ^69^ and Dr Sian Morgan ^9^.

**Metadata curation:**

Dr Elizabeth Wastenge ^53^, Dr Michael R Chapman ^34, 88, 99^, Mr Siddharth Mookerjee ^38, 103^, Dr Rachael Stanley ^54^, Mrs Wendy Smith ^15^, Prof Timothy Peto ^59^, Dr David Eyre ^59^, Dr Derrick Crook ^59^, Dr Gabrielle Vernet ^33^, Dr Christine Kitchen ^10^, Huw Gulliver ^10^, Dr Ian Merrick ^10^, Prof Martyn Guest ^10^, Robert Munn ^10^, Dr Declan T Bradley ^63, 72^, and Dr Tim Wyatt ^63^.

**Project administration:**

Dr Charlotte Beaver ^99^, Luke Foulser ^99^, Sophie Palmer ^88^, Carol M Churcher ^88^, Ellena Brooks ^88^, Kim S Smith ^88^, Dr Katerina Galai ^88^, Georgina M McManus ^88^, Dr Frances Bolt ^38, 103^, Dr Francesc Coll ^19^, Lizzie Meadows ^70^, Dr Stephen W Attwood ^23^, Dr Alisha Davies ^69^, Elen De Lacy ^69^, Fatima Downing ^69^, Sue Edwards ^69^, Dr Garry P Scarlett ^76^, Mrs Sarah Jeremiah ^83^ and Dr Nikki Smith ^93^.

**Samples and logistics:**

Danielle Leek ^88^, Sushmita Sridhar ^88, 99^, Sally Forrest ^88^, Claire Cormie ^88^, Harmeet K Gill ^88^, Joana Dias ^88^, Ellen E Higginson ^88^, Mailis Maes ^88^, Jamie Young ^88^, Michelle Wantoch ^7^, Sanger Covid Team (www.sanger.ac.uk/covid-team) ^99^, Dorota Jamrozy ^99^, Stephanie Lo ^99^, Dr Minal Patel ^99^, Verity Hill ^90^, Ms Claire M Bewshea ^91^, Prof Sian Ellard ^73, 91^, Dr Cressida Auckland ^73^, Dr Ian Harrison ^66^, Dr Chloe Bishop ^66^, Dr Vicki Chalker ^66^, Dr Alex Richter ^85^, Dr Andrew Beggs ^85^, Dr Angus Best ^86^, Dr Benita Percival ^86^, Dr Jeremy Mirza ^86^, Dr Oliver Megram ^86^, Dr Megan Mayhew ^86^, Dr Liam Crawford ^86^, Dr Fiona Ashcroft ^86^, Dr Emma Moles-Garcia ^86^, Dr Nicola Cumley ^86^, Mr Richard Hopes ^64^, Dr Patawee Asamaphan ^48^, Mr Marc O Niebel ^48^, Prof Rory N Gunson ^100^, Dr Amanda Bradley ^52^, Dr Alasdair Maclean ^52^, Dr Guy Mollett ^52^, Dr Rachel Blacow ^52^, Mr Paul Bird ^16^, Mr Thomas Helmer ^16^, Miss Karlie Fallon ^16^, Dr Julian Tang ^16^, Dr Antony D Hale ^49^, Dr Louissa R Macfarlane-Smith ^49^, Katherine L Harper ^49^, Miss Holli Carden ^49^, Dr Nicholas W Machin ^45, 64^, Ms Kathryn A Jackson ^92^, Dr Shazaad S Y Ahmad ^45, 64^, Dr Ryan P George ^45^, Dr Lance Turtle ^92^, Mrs Elaine O’Toole ^43^, Mrs Joanne Watts ^43^, Mrs Cassie Breen ^43^, Mrs Angela Cowell ^43^, Ms Adela Alcolea-Medina ^32, 96^, Ms Themoula Charalampous ^12, 42^, Amita Patel ^11^, Dr Lisa J Levett ^35^, Dr Judith Heaney ^35^, Dr Aileen Rowan ^39^, Prof Graham P Taylor ^39^, Dr Divya Shah ^30^, Miss Laura Atkinson ^30^, Mr Jack CD Lee ^30^, Mr Adam P Westhorpe ^82^, Dr Riaz Jannoo ^82^, Dr Helen L Lowe ^82^, Miss Angeliki Karamani ^82^, Miss Leah Ensell ^82^, Mrs Wendy Chatterton ^35^, Miss Monika Pusok ^35^, Mrs Ashok Dadrah ^75^, Miss Amanda Symmonds ^75^, Dr Graciela Sluga ^44^, Dr Zoltan Molnar ^72^, Mr Paul Baker ^79^, Prof Stephen Bonner ^79^, Ms Sarah Essex ^79^, Dr Edward Barton ^56^, Ms Debra Padgett ^56^, Ms Garren Scott ^56^, Ms Jane Greenaway ^57^, Dr Brendan AI Payne ^50^, Dr Shirelle Burton-Fanning ^50^, Dr Sheila Waugh ^50^, Dr Veena Raviprakash ^17^, Ms Nicola Sheriff ^17^, Ms Victoria Blakey ^17^, ms Lesley-Anne Williams ^17^, Dr Jonathan Moore ^27^, Ms Susanne Stonehouse ^27^, Dr Louise Smith ^55^, Dr Rose K Davidson ^89^, Dr Luke Bedford ^26^, Dr Lindsay Coupland ^54^, Ms Victoria Wright ^18^, Dr Joseph G Chappell ^97^, Dr Theocharis Tsoleridis ^97^, Prof Jonathan Ball ^97^, Mrs Manjinder Khakh ^15^, Dr Vicki M Fleming ^15^, Dr Michelle M Lister ^15^, Dr Hannah C Howson-Wells ^15^, Dr Louise Berry ^15^, Dr Tim Boswell ^15^, Dr Amelia Joseph ^15^, Dr Iona Willingham ^15^, Dr Nichola Duckworth ^60^, Dr Sarah Walsh ^60^, Dr Emma Wise ^2, 111^, Dr Nathan Moore ^2, 111^, Miss Matilde Mori ^2, 108, 111^, Dr Nick Cortes ^2, 111^, Dr Stephen Kidd ^2, 111^, Dr Rebecca Williams ^33^, Laura Gifford ^69^, Miss Kelly Bicknell ^61^, Dr Sarah Wyllie ^61^, Miss Allyson Lloyd ^61^, Mr Robert Impey ^61^, Ms Cassandra S Malone ^6^, Mr Benjamin J Cogger ^6^, Nick Levene ^62^, Lynn Monaghan ^62^, Dr Alexander J Keeley ^93^, Dr David G Partridge ^78, 93^, Dr Mohammad Raza ^78, 93^, Dr Cariad Evans ^78, 93^ and Dr Kate Johnson ^78, 93^.

**Sequencing and analysis:**

Emma Betteridge ^99^, Ben W Farr ^99^, Scott Goodwin ^99^, Dr Michael A Quail ^99^, Carol Scott ^99^, Lesley Shirley ^99^, Scott AJ Thurston ^99^, Diana Rajan ^99^, Dr Iraad F Bronner ^99^, Louise Aigrain ^99^, Dr Nicholas M Redshaw ^99^, Dr Stefanie V Lensing ^99^, Shane McCarthy ^99^, Alex Makunin ^99^, Dr Carlos E Balcazar ^90^, Dr Michael D Gallagher ^90^, Dr Kathleen A Williamson ^90^, Thomas D Stanton ^90^, Ms Michelle L Michelsen ^91^, Ms Joanna Warwick-Dugdale ^91^, Dr Robin Manley ^91^, Ms Audrey Farbos ^91^, Dr James W Harrison ^91^, Dr Christine M Sambles ^91^, Dr David J Studholme ^91^, Dr Angie Lackenby ^66^, Dr Tamyo Mbisa ^66^, Dr Steven Platt ^66^, Mr Shahjahan Miah ^66^, Dr David Bibby ^66^, Dr Carmen Manso ^66^, Dr Jonathan Hubb ^66^, Dr Gavin Dabrera ^66^, Dr Mary Ramsay ^66^, Dr Daniel Bradshaw ^66^, Dr Ulf Schaefer ^66^, Dr Natalie Groves ^66^, Dr Eileen Gallagher ^66^, Dr David Lee ^66^, Dr David Williams ^66^, Dr Nicholas Ellaby ^66^, Hassan Hartman ^66^, Nikos Manesis ^66^, Vineet Patel ^66^, Juan Ledesma ^67^, Ms Katherine A Twohig ^67^, Dr Elias Allara ^64, 88^, Ms Clare Pearson ^64, 88^, Mr Jeffrey K. J. Cheng ^94^, Dr Hannah E. Bridgewater ^94^, Ms Lucy R. Frost ^94^, Ms Grace Taylor-Joyce ^94^, Dr Paul E Brown ^94^, Dr Lily Tong ^48^, Ms Alice Broos ^48^, Mr Daniel Mair ^48^, Mrs Jenna Nichols ^48^, Dr Stephen N Carmichael ^48^, Dr Katherine L Smollett ^40^, Dr Kyriaki Nomikou ^48^, Dr Elihu Aranday-Cortes ^48^, Ms Natasha Johnson ^48^, Dr Seema Nickbakhsh ^48, 68^, Dr Edith E Vamos ^92^, Dr Margaret Hughes ^92^, Dr Lucille Rainbow ^92^, Mr Richard Eccles ^92^, Ms Charlotte Nelson ^92^, Dr Mark Whitehead ^92^, Dr Richard Gregory ^92^, Mr Matthew Gemmell ^92^, Ms Claudia Wierzbicki ^92^, Ms Hermione J Webster ^92^, Ms Chloe L Fisher ^28^, Mr Adrian W Signell ^20^, Dr Gilberto Betancor ^20^, Mr Harry D Wilson ^20^, Dr Gaia Nebbia ^12^, Dr Flavia Flaviani ^31^, Mr Alberto C Cerda ^96^, Ms Tammy V Merrill ^96^, Rebekah E Wilson ^96^, Mr Marius Cotic ^82^, Miss Nadua Bayzid ^82^, Dr Thomas Thompson ^72^, Dr Erwan Acheson ^72^, Prof Steven Rushton ^51^, Prof Sarah O’Brien ^51^, David J Baker ^70^, Steven Rudder ^70^, Alp Aydin ^70^, Dr Fei Sang ^18^, Dr Johnny Debebe ^18^, Dr Sarah Francois ^23^, Dr Tetyana I Vasylyeva ^23^, Dr Marina Escalera Zamudio ^23^, Mr Bernardo Gutierrez ^23^, Dr Angela Marchbank ^10^, Joshua Maksimovic ^9^, Karla Spellman ^9^, Kathryn McCluggage ^9^, Dr Mari Morgan ^69^, Robert Beer ^9^, Safiah Afifi ^9^, Trudy Workman ^10^, William Fuller ^10^, Catherine Bresner ^10^, Dr Adrienn Angyal ^93^, Dr Luke R Green ^93^, Dr Paul J Parsons ^93^, Miss Rachel M Tucker ^93^, Dr Rebecca Brown ^93^ and Mr Max Whiteley ^93^.

**Software and analysis tools:**

James Bonfield ^99^, Dr Christoph Puethe ^99^, Mr Andrew Whitwham ^99^, Jennifier Liddle ^99^, Dr Will Rowe ^41^, Dr Igor Siveroni ^39^, Dr Thanh Le-Viet ^70^ and Amy Gaskin ^69^.

**Visualisation:**

Dr Rob Johnson ^39^.

**1** Barking, Havering and Redbridge University Hospitals NHS Trust, **2** Basingstoke Hospital, **3** Belfast Health & Social Care Trust, **4** Betsi Cadwaladr University Health Board, **5** Big Data Institute, Nuffield Department of Medicine, University of Oxford, **6** Brighton and Sussex University Hospitals NHS Trust, **7** Cambridge Stem Cell Institute, University of Cambridge, **8** Cambridge University Hospitals NHS Foundation Trust, **9** Cardiff and Vale University Health Board, **10** Cardiff University, **11** Centre for Clinical Infection & Diagnostics Research, St. Thomas’ Hospital and Kings College London, **12** Centre for Clinical Infection and Diagnostics Research, Department of Infectious Diseases, Guy’s and St Thomas’ NHS Foundation Trust, **13** Centre for Enzyme Innovation, University of Portsmouth (PORT), **14** Centre for Genomic Pathogen Surveillance, University of Oxford, **15** Clinical Microbiology Department, Queens Medical Centre, **16** Clinical Microbiology, University Hospitals of Leicester NHS Trust, **17** County Durham and Darlington NHS Foundation Trust, **18** Deep Seq, School of Life Sciences, Queens Medical Centre, University of Nottingham, **19** Department of Infection Biology, Faculty of Infectious & Tropical Diseases, London School of Hygiene & Tropical Medicine, **20** Department of Infectious Diseases, King’s College London, **21** Department of Microbiology, Kettering General Hospital, **22** Departments of Infectious Diseases and Microbiology, Cambridge University Hospitals NHS Foundation Trust; Cambridge, UK, **23** Department of Zoology, University of Oxford, **24** Division of Virology, Department of Pathology, University of Cambridge, **25** East Kent Hospitals University NHS Foundation Trust, **26** East Suffolk and North Essex NHS Foundation Trust, **27** Gateshead Health NHS Foundation Trust, **28** Genomics Innovation Unit, Guy’s and St. Thomas’ NHS Foundation Trust, **29** Gloucestershire Hospitals NHS Foundation Trust, **30** Great Ormond Street Hospital for Children NHS Foundation Trust, **31** Guy’s and St. Thomas’ BRC, **32** Guy’s and St. Thomas’ Hospitals, **33** Hampshire Hospitals NHS Foundation Trust, **34** Health Data Research UK Cambridge, **35** Health Services Laboratories, **36** Heartlands Hospital, Birmingham, **37** Hub for Biotechnology in the Built Environment, Northumbria University, **38** Imperial College Hospitals NHS Trust, **39** Imperial College London, **40** Institute of Biodiversity, Animal Health & Comparative Medicine, **41** Institute of Microbiology and Infection, University of Birmingham, **42** King’s College London, **43** Liverpool Clinical Laboratories, **44** Maidstone and Tunbridge Wells NHS Trust, **45** Manchester University NHS Foundation Trust, **46** Microbiology Department, Wye Valley NHS Trust, Hereford, **47** MRC Biostatistics Unit, University of Cambridge, **48** MRC-University of Glasgow Centre for Virus Research, **49** National Infection Service, PHE and Leeds Teaching Hospitals Trust, **50** Newcastle Hospitals NHS Foundation Trust, **51** Newcastle University, **52** NHS Greater Glasgow and Clyde, **53** NHS Lothian, **54** Norfolk and Norwich University Hospital, **55** Norfolk County Council, **56** North Cumbria Integrated Care NHS Foundation Trust, **57** North Tees and Hartlepool NHS Foundation Trust, **58** Northumbria University, **59** Oxford University Hospitals NHS Foundation Trust, **60** PathLinks, Northern Lincolnshire & Goole NHS Foundation Trust, **61** Portsmouth Hospitals University NHS Trust, **62** Princess Alexandra Hospital Microbiology Dept., **63** Public Health Agency, **64** Public Health England, **65** Public Health England, Clinical Microbiology and Public Health Laboratory, Cambridge, UK, **66** Public Health England, Colindale, **67** Public Health England, Colindale, **68** Public Health Scotland, **69** Public Health Wales NHS Trust, **70** Quadram Institute Bioscience, **71** Queen Elizabeth Hospital, **72** Queen’s University Belfast, **73** Royal Devon and Exeter NHS Foundation Trust, **74** Royal Free NHS Trust, **75** Sandwell and West Birmingham NHS Trust, **76** School of Biological Sciences, University of Portsmouth (PORT), **77** School of Pharmacy and Biomedical Sciences, University of Portsmouth (PORT), **78** Sheffield Teaching Hospitals, **79** South Tees Hospitals NHS Foundation Trust, **80** Swansea University, **81** University Hospitals Southampton NHS Foundation Trust, **82** University College London, **83** University Hospital Southampton NHS Foundation Trust, **84** University Hospitals Coventry and Warwickshire, **85** University of Birmingham, **86** University of Birmingham Turnkey Laboratory, **87** University of Brighton, **88** University of Cambridge, **89** University of East Anglia, **90** University of Edinburgh, **91** University of Exeter, **92** University of Liverpool, **93** University of Sheffield, **94** University of Warwick, **95** University of Cambridge, **96** Viapath, Guy’s and St Thomas’ NHS Foundation Trust, and King’s College Hospital NHS Foundation Trust, **97** Virology, School of Life Sciences, Queens Medical Centre, University of Nottingham, **98** Wellcome Centre for Human Genetics, Nuffield Department of Medicine, University of Oxford, **99** Wellcome Sanger Institute, **100** West of Scotland Specialist Virology Centre, NHS Greater Glasgow and Clyde, **101** Department of Medicine, University of Cambridge, **102** Ministry of Health, Sri Lanka, **103** NIHR Health Protection Research Unit in HCAI and AMR, Imperial College London, **104** North West London Pathology, **105** NU-OMICS, Northumbria University, **106** University of Kent, **107** University of Oxford, **108** University of Southampton, **109** University of Southampton School of Health Sciences, **110** University of Southampton School of Medicine, **111** University of Surrey, **112** Warwick Medical School and Institute of Precision Diagnostics, Pathology, UHCW NHS Trust, **113** Wellcome Africa Health Research Institute Durban and **114** Wellcome Genome Campus.

## Footnotes

1 We calculated this as the posterior mean and 2.5-97.5 quantiles of the set of 42×6 posterior medians of the distributions (R_{S-} - R_{S+}), one per STP-week

